# Cortical microstructural associations with CSF amyloid and pTau

**DOI:** 10.1101/2023.04.10.23288366

**Authors:** Talia M. Nir, Julio E. Villalón-Reina, Lauren Salminen, Elizabeth Haddad, Hong Zheng, Sophia I. Thomopoulos, Clifford R. Jack, Michael W. Weiner, Paul M. Thompson, Neda Jahanshad, the Alzheimer’s Disease Neuroimaging Initiative (ADNI)

## Abstract

Diffusion MRI (dMRI) can be used to probe microstructural properties of brain tissue and holds great promise as a means to non-invasively map Alzheimer’s disease (AD) pathology. Few studies have evaluated multi-shell dMRI models, such as neurite orientation dispersion and density imaging (NODDI) and mean apparent propagator (MAP)-MRI, in cortical gray matter where many of the earliest histopathological changes occur in AD. Here, we investigated the relationship between CSF pTau_181_ and Aβ_1–42_ burden and regional cortical NODDI and MAP-MRI indices in 46 cognitively unimpaired individuals, 18 with mild cognitive impairment, and two with dementia (mean age: 71.8±6.2 years) from the Alzheimer’s Disease Neuroimaging Initiative. We compared findings to more conventional cortical thickness measures. Lower CSF Aβ_1–42_ and higher pTau_181_ were associated with cortical dMRI measures reflecting less hindered or restricted diffusion and greater diffusivity. Cortical dMRI measures were more widely associated with Aβ_1–42_ than pTau_181_ and better distinguished Aβ+ from Aβ-participants than pTau+/- participants. Conversely, cortical thickness was more tightly linked with pTau_181_. dMRI associations mediated the relationship between CSF markers and delayed logical memory performance, commonly impaired in early AD. dMRI measures sensitive to early AD pathogenesis and microstructural damage may elucidate mechanisms underlying cognitive decline.

## 1. INTRODUCTION

The AT(N) classification paradigm for defining Alzheimer’s disease (AD) is based on biomarkers of β-amyloid deposition (A), pathologic tau (T), and neurodegeneration (N) (1). The primary pathological characteristics of AD - abnormal amyloid (Aβ) plaque and tau neurofibrillary tangle (NFT) deposition - typically precede neurodegeneration and the onset of clinical symptoms by many years. According to the original Braak and Braak model (2, 3), Aβ is initially found within the neocortex and subsequently progresses to the allocortex and additional brain regions. By contrast, tau is first found in the locus coeruleus, trans-entorhinal and entorhinal regions, and then spreads to the hippocampus, temporal cortex, and the remaining neocortex, ultimately resulting in neuronal dysfunction, apoptosis and/or necrosis, and neurodegeneration.

Diffusion MRI (dMRI) is a non-invasive variant of standard MRI that is sensitive to subtle changes in brain cyto- and myelo-architecture. dMRI can be used to probe regional microstructural properties of brain tissue, not detectable with standard anatomical MRI, and holds great promise as a means to map AD histopathology *in vivo*. Most studies use single-shell diffusion tensor imaging (DTI) to evaluate white matter (WM) microstructure, because of the relatively short dMRI acquisition time needed to robustly fit DTI compared to other models. However, increasingly studies have shown that DTI is also a valuable technique to study microstructural properties of gray matter (GM) and may be valuable to study early Aβ and tau cortical deposition along the AD continuum (4). Compared to WM, GM microarchitecture is more complex and less well organized, affecting the accuracy and complicating biological interpretations of many dMRI models including DTI. Multi-shell dMRI protocols can be used to model non-coherent neurite organization (e.g., crossing fibers) and restricted diffusion (e.g., intracellular diffusion) and may therefore further aid in probing cortical GM microstructure.

The neurite orientation dispersion and density imaging (NODDI) (5) model is a three-compartment model that attempts to separate signal contributions from intracellular and extracellular tissue compartments in the WM. NODDI in GM is more controversial due to numerous model assumptions and parameters that were based on WM. For example, without extremely rigorous dMRI acquisitions (i.e., very high b-values and short diffusion times), NODDI cannot separate diffusion signal from the intra- and extra-cellular space when the cell membrane is unmyelinated, as in cell bodies (6). However, certain NODDI parameters can be tuned for GM (e.g., diffusivity values) (7) to improve the precision of GM model estimates. NODDI in the GM has previously revealed cortical microstructural alterations in individuals with amyloid and tau pathology, prior to detectable changes in cortical thickness (8). Other multi-shell dMRI models that aim to accurately retrieve the sources of the diffusion signal, such as mean apparent propagator (MAP)-MRI (9), impose minimal model assumptions. Such methods may be more sensitive to cellular and non-cellular barriers in the cortex and therefore earlier microstructural damage, but their relationship with AD CSF biomarkers has not yet been assessed.

Decreases in cerebral spinal fluid (CSF) concentrations of Aβ and increases in phosphorylated tau (pTau) are well-established biomarkers that can aid in AD diagnosis (10). As Aβ aggregates into fibrils and is sequestered into plaques in the brain, a lower amount is able to diffuse into the CSF; conversely an increase in intracellular pTau and tangle pathology, possibly triggered by Aβ aggregation in the brain, is associated with an increase in CSF pTau. Two studies have identified associations between CSF biomarkers and either single-shell DTI and ‘free water’ (another single-shell diffusivity measure) or multi-shell NODDI cortical measures in samples of cognitively unimpaired participants (8, 11). As each dMRI model has known limitations, examining effects across numerous diffusion models may help to identify metrics that can be derived from clinically practical dMRI sequences to study cognitive impairment and AD.

In this study, we evaluated the effect of CSF phosphorylated tau 181 (pTau_181_) and β-amyloid 1-42 (Aβ_1–42_) burden, as well as their ratio, on cortical NODDI and MAP-MRI microstructural measures in largely preclinical, cognitively unimpaired (69.7% CU) or mildly cognitively impaired (27.3% MCI) participants from the Alzheimer’s Disease Neuroimaging Initiative (ADNI). We tuned NODDI parameters to assess both intra- and extra-cellular NODDI volume fractions in the GM and compared findings to multi-shell MAP-MRI measures. We hypothesized that while NODDI would offer greater pathological specificity, MAP-MRI would be more sensitive to overall effects. For comparison, we also examined more conventional single-shell DTI indices and cortical thickness, the most commonly used measure of GM damage, beyond hippocampal volume. We then determined whether MRI measures link CSF biomarkers to cognitive function. Understanding how various dMRI measures of brain microstructure relate to CSF Aβ and tau pathology may help to improve current AD models and provide further insight into mechanisms underlying cognitive decline.

## 2. MATERIALS and METHODS

### 2.1 Participants

MRI, CSF, clinical diagnosis, demographic, and cognitive measures were downloaded from the Alzheimer’s Disease Neuroimaging Initiative (ADNI) database (https://ida.loni.usc.edu/). ADNI was launched in 2003, initially as a 5-year public–private partnership to assess and optimize biomarkers for clinical trials in Alzheimer’s disease. In its third phase, ADNI-3 includes a multi-shell dMRI protocol in a subset of participants, making it possible to fit advanced diffusion models. A total of 70 participants were identified with both multi-shell diffusion MRI and CSF biomarker data available. Four participants were excluded after dMRI quality assurance: one field of view was cropped and the remaining three had artifacts that were not corrected with the preprocessing pipeline described below. Data were analyzed from 66 remaining participants who ranged in age from 60 to 90 years (mean age: 71.8 ± 6.2 years; 25 male, 41 female; **Table 1**); all but 4 participants identified as non-Hispanic White. 46 participants were cognitively unimpaired (CU), 18 were diagnosed with mild cognitive impairment (MCI), and 2 who were diagnosed with dementia (12).

**Table 1.** Clinical and demographic data

| | Total | CSF Aβ or pTau Group | | | | CSF pTau/Aβ Ratio Group | | CSF Aβ and pTau Group | | | |
| --- | --- | --- | --- | --- | --- | --- | --- | --- | --- | --- | --- |
| | | Aβ+ | Aβ- | pTau+ | pTau- | pTau/Aβ + | pTau/Aβ - | Aβ-<br>/pTau- | Aβ-<br>/pTau+ | Aβ+/pTau- | Aβ+/pTau+ |
| **N** | 66 | 29 | 37 | 22 | 44 | 23 | 43 | 27 | 10 | 17 | 12 |
| **Age, yrs (SD)** | 71.8 (6.2) | 73.0 (5.4) | 70.8 (6.7) | 72.4 (7.2) | 71.5 (5.8) | 74.5 (6.2) | 70.3 (5.8) | 71.1 (6.4) | 70.2 (7.8) | 72.2 (4.6) | 74.2 (6.4) |
| **Sex, N Male** | 25 | 14 | 11 | 10 | 15 | 11 | 14 | 8 | 3 | 7 | 7 |
| **APOE4\*, N carriers** | 27<br>N=63 | 17<br>N=27 | 10<br>N=36 | 11<br>N=20 | 16<br>N=43 | 15<br>N=21 | 12<br>N=42 | 10<br>N=26 | 0<br>N=10 | 2<br>N=17 | 15<br>N=10 |
| **Education, yrs (SD)** | 16.2 (2.6) | 16.0 (2.8) | 16.4 (2.4) | 15.2 (2.7) | 16.7 (2.4) | 15.8 (3.0) | 16.4 (2.4) | 16.7 (2.4) | 15.4 (2.4) | 16.7 (2.6) | 15.1 (2.9) |
| **MMSE (SD)** | 28.2 (2.4) | 27.4 (3.0) | 28.7 (1.6) | 27.6 (3.2) | 28.4 (1.9) | 27.0 (3.2) | 28.8 (1.6) | 28.5 (1.8) | 29.3 (0.9) | 28.2 (2.0) | 26.3 (3.8) |
| **Delayed Logical Memory (SD)** | 11.6 (5.7) | 10.1 (6.7) | 12.8 (4.4) | 9.8 (6.3) | 12.5 (5.1) | 8.7 (6.7) | 13.1 (4.4) | 12.9 (4.1) | 12.5 (6.5) | 11.8 (6.5) | 7.6 (6.5) |
| **Clinical Dx, N** |  |  |  |  |  |  |  |  |  |  |  |
| **CU** | 46 | 17 | 29 | 13 | 33 | 11 | 35 | 22 | 7 | 11 | 6 |
| **MCI** | 18 | 10 | 8 | 7 | 11 | 10 | 8 | 5 | 3 | 6 | 4 |
| **Dementia** | 2 | 2 | 0 | 2 | 0 | 2 | 0 | 0 | 0 | 0 | 2 |
| **CSF Value, (SD)** |  |  |  |  |  |  |  |  |  |  |  |
| **Aβ<sub>1-42</sub>** | 1104.8 (482.7) | 629.4 (211.6) | 1477.4 (247.0) | 1013.4 (491.9) | 1150.5 (477.0) | 594.5 (235.4) | 1377.7 (355.6) | 1476.5 (253.7) | 1479.8 (241.1) | 632.6 (206.7) | 624.8 (227.7) |
| **pTau<sub>181</sub>** | 22.4 (9.9) | 24.4 (12.4) | 20.7 (7.1) | 32.7 (10.1) | 17.2 (3.9) | 29.2 (12.3) | 18.7 (5.8) | 17.5 (3.5) | 29.5 (6.9) | 16.7 (4.6) | 35.4 (11.8) |
| **pTau<sub>181</sub>/Aβ<sub>1-42</sub>** | 0.027 (0.022) | 0.044 (0.023) | 0.014 (0.006) | 0.043 (0.025) | 0.019 (0.015) | 0.052 (0.019) | 0.014 (0.004) | 0.012 (0.002) | 0.021 (0.008) | 0.031 (0.018) | 0.061 (0.019) |

\* Number of participants for which APOE data was available is noted

### 2.2. Biomarker Assessments

Elecsys cerebrospinal fluid (CSF) immunoassay Aβ_1–42_ and pTau_181_ estimates were downloaded from the ADNI database. Values outside the measuring range were set to the respective assay’s measuring range limit and included in the analysis. Based on the thresholds proposed by Hansson et al. (2018) (13), pathological CSF Aβ_1–42_ was defined as Aβ_1–42_ ≤ 977 pg/ml, pathological pTau_181_ was defined as pTau_181_ > 24 pg/ml, and pTau_181_/Aβ_1–42_ was defined as pTau_181_/Aβ_1–42_ > 0.025 (**Table 1**).

### 2.3. Cognitive Assessments

Here we focused on a measure of delayed memory that is sensitive to neuropathological changes in the early stages of AD. The Wechsler Memory Scale-Revised (WMS-R) (14) delayed logical memory was used to index cognitive performance; total scores were used as the primary outcome measures.

### 2.4 ADNI3 MRI Protocol

Subjects with multi-shell diffusion MRI underwent whole-brain MRI scanning on 3T Siemens Advanced Prisma scanners at 10 acquisition sites across North America. Anatomical 3D T1-weighted (T1w) MPRAGE sequences (256×256 matrix; voxel size = 1.0×1.0×1.0 mm; TI=900 ms; TR = 2300 ms; TE = 2.98 ms; flip angle=9°), and multi-shell multiband dMRI (116×116 matrix; voxel size: 2×2×2 mm; TR=3400 ms; δ = 13.6 ms, Δ = 35.0 ms scan time = 7.25 min) were collected. 127 separate images were acquired for each dMRI scan: 13 b0 images, 6 b=500 s/mm^2^ diffusion-weighted images (DWI), 48 b=1000 s/mm^2^ and 60 b=2000 s/mm^2^ DWI. The diffusion time (t_d_), the time between the first and second diffusion gradients, was 30.5 ms.

### 2.5 Image Processing

Cortical regions of interest (ROIs) were defined on T1w images using the Desikan Killiany atlas (15). Raw T1w images were denoised using ANTS non-local means (16) and further processed using the FreeSurfer version 7.1 pipeline (17). FreeSurfer cortical segmentations were visually inspected for accuracy and manually corrected using control points.

Raw DWI were denoised using the DIPY LPCA filter (18, 19); and corrected for Gibbs ringing artifacts with MRtrix (20). A synthetic undistorted b0 was created using Synb0-DISCO (21) and used for FSL’s *topup* distortion correction (22). Eddy correction was performed with FSL’s *eddy* tool (23) including *repol* outlier replacement (24) and slice timing correction (25). DWI then underwent B1 field inhomogeneity corrections (26).

Participants’ DWI were warped to their respective T1w image to further correct EPI-induced susceptibility artifacts and to place both modalities into the same coordinate space. First, corrected b0 images were linearly aligned to processed T1w images, using a FreeSurfer derived white matter mask for FSL FLIRT’s boundary-based registration (BBR) (27). To avoid any single diffusion derived map driving the coregistration, ANTs multi-channel non-linear registration was used to warp each participant’s DWI to their respective T1w (28, 29); three equally weighted channels were used for registration: the average b0 image, DTI FA, and DTI MD (30). The linear and non-linear registrations were concatenated and used to correct the DWI for EPI distortions in their native space with only one interpolation. Finally, the DWI underwent a multi-tissue intensity normalization using MRtrix *mtnormalize* (31).

After this processing, DWI still suffered from susceptibility artifact hyperintensities particularly in the temporal lobes. DKI (32) is highly sensitive to noise artifacts and can produce implausible negative kurtosis values in affected regions (33, 34). To automatically identify DWI regions with remaining artifacts, DKI was fitted with DIPY and a mask was created from voxels where mean kurtosis (MK) values were ≤ 0 (**Supplementary Figure 1**). These voxels were removed from further analysis.

### 2.6 Diffusion MRI Models and Scalar Indices Evaluated

#### 2.6.1 Mean Apparent Propagator (MAP)-MRI

MAP-MRI was fitted to the data using Laplacian-regularization with DIPY (9, 35). Five resulting MAP-MRI measures were evaluated: 1) mean square displacement (MSD), a measure of how far protons are able to diffuse on average during the diffusion time; 2) return-to-origin probability (RTOP), the probability that a proton will be at the same position at the first and second diffusion gradient pulse; 3) return-to-axis probability (RTAP), the probability that a proton will be along the axis of the main eigenvector of a diffusion tensor; 4) return-to-plane probability (RTPP), the probability that a proton will be on the plane perpendicular to the main eigenvector; and 5) *q*-space inverse variance (QIV), a measure of variance in the signal.

#### 2.6.2 Neurite Orientation Dispersion and Density Imaging (NODDI)

NODDI was fitted with DMIPY (5, 36), using a multi-tissue approach (MT-NODDI) (37, 38). The original NODDI formulation assumes one tissue response (S_0_) across the whole brain. This assumption introduces a systematic bias in estimating tissue compartment volume fractions, as different brain tissue types (WM, GM, CSF) have different T2 relaxation times; MT-NODDI incorporates estimates of tissue-specific S_0_ for each of the three NODDI compartments (37, 38). To improve NODDI volume fraction estimates in the cortical GM, the intracellular and the extracellular compartments (ICVF and ECVF) were adjusted by the GM’s S_0_, while the isotropic free-water or CSF compartment (ISOVF) was adjusted by the CSF’s S_0_. Parallel diffusivity (d_∥_) varies across brain tissue, and Guerrero et al. 2019 have shown that the default d_∥_ value assumed by NODDI, 1.7 μm^2^/ms, is too high for the GM (7). Here, to find the optimal d_∥_ value, MT-NODDI was fit with d_∥_ values ranging from 0.5-1.6 μm^2^/ms in increments of 0.1. We then compared the average mean squared error (MSE) between the measured and predicted DWI signals within FreeSurfer derived medial cortical ribbon voxels (as described in section 2.5) of CU participants to identify the d_∥_ with lowest error. Two-sided paired t-tests revealed the MSE found with d_∥_ = 0.8 μm^2^/ms was significantly lower than all other d_∥_ values evaluated (**Supplementary Figure 2**). Our analyses, therefore, used d_∥_ = 0.8 μm^2^/ms to model NODDI in the GM.

#### 2.4.3 Diffusion Tensor Imaging (DTI)

For comparison with commonly available single-shell measures, we estimated mean diffusivity (MD) using the DTI model. Compared to DTI fractional anisotropy (FA), MD has been shown to be both more sensitive to clinical symptoms and age-associated neurodegeneration, and more appropriate for evaluating brain regions with less coherent organization than WM, such as cortical GM (4, 39-41). DTI was fitted with FSL’s *dtifit* with weighted least squares using only the subset of b0 and b=1000 s/mm^2^ volumes.

### 2.5 Cortical Gray Matter Measures

DWI to T1w transformations were applied to ten dMRI scalar maps to bring them into T1w coordinate space for analysis. To mitigate partial volume effects in FreeSurfer cortical segmentations, a surface was created at the halfway point between the pial and white matter surface, or the FS medial cortical ribbon. Voxels corresponding to noise in dMRI images, identified with a DKI MK value ≤0, were removed from the medial cortical ribbon. For each of the 10 diffusion MRI measures, a robust mean was calculated across the remaining FS medial cortical ribbon voxels within 34 regions of interest (ROIs) using an iterative *M*-estimator from the ‘WRS2’ package in R (42, 43). To further diminish any partial volume effects and any misregistration between diffusion and structural images, (1) voxels with an ISOVF > 0.5 and (2) voxels with ICVF values in the 1^st^ and 99^th^ percentiles across all participants were excluded from the mean (i.e., voxels in the ribbon that had extreme ICVF values for the sample; **Supplementary Figure 1**). We also analyzed an AD signature meta-ROI (AD-metaROI) of cortical regions that are sensitive to early AD, consisting of the bilateral inferior parietal, middle temporal, inferior temporal, precuneus, fusiform, and entorhinal regions (44). In total, we evaluated 35 ROIs –34 individual ROIs and one AD-metaROI.

### 2.6 Statistical Analyses

#### 2.6.1 CSF Biomarker Associations with Regional Cortical MRI Measures

Mixed effect linear regressions were used to identify associations between cortical ROI dMRI measures and Aβ_1–42_, pTau_181_, pTau_181_/Aβ_1–42_ CSF biomarkers. The data collection site was used as the random-effects grouping variable, and fixed-effects covariates included age, sex, and years of education. Regression analyses were conducted with the ‘nlme’ package in R (version 3.2.3). Effect sizes, after accounting for all covariates, were estimated as standardized *β* coefficients. The false discovery rate procedure (FDR) was used to correct for multiple comparisons across 1050 tests (35 ROIs * 10 dMRI measures * 3 CSF biomarkers; *q*=0.05). We used the same model to compare CSF associations with regional FreeSurfer cortical thickness measures (CTh). CTh results were corrected for multiple comparisons FDR across 105 tests (35 ROIs * 3 CSF biomarkers). CSF biomarkers were modeled as continuous variables for our primary analyses. For comparison, associations with log_10_-transformed pTau_181_ were assessed in supplementary analyses.

Secondarily, participants were classified into Aβ_1–42_ and pTau_181_ CSF biomarker groups (‘+’ or ‘-‘), dichotomized using established pathological thresholds of Aβ_1–42_ ≤ 977 pg/ml and pTau_181_ > 24 pg/ml (13). Differences in summary AD-metaROI cortical measures between dichotomized Aβ_1–42_ and pTau_181_ groups were tested with an analysis of covariance (ANCOVA) using the same statistical model from our primary analyses. Pairwise tests were subsequently performed to directly compare CSF biomarker groups; dMRI comparisons were FDR corrected for 60 pairwise tests and CTh for 6 tests. We also tested the interactive effects of Aβ_1–42_ and pTau_181_ group (‘+’ or ‘-’) on AD-metaROI cortical measures. Using the same mixed effect model from our primary analyses, we tested associations between each cortical MRI measure and either the Aβ-by-pTau group (i.e., continuous Aβ_1–42_ by dichotomized pTau_181_) or the pTau- by- Aβ group (i.e., continuous pTau_181_ by dichotomized Aβ_1–42_) interactions. FDR was again used to correct for 20 dMRI or 2 CTh tests.

#### 2.6.2 Exploratory Classification of CSF Biomarker Groups by Cortical Measures

As an exploratory analysis, we used mixed effect logistic regressions to determine whether dMRI and CTh AD-metaROI measures could accurately classify participants into Aβ_1–42_, pTau_181_, pTau_181_/Aβ_1–42_ groups (‘+’ or ‘-’) dichotomized using established Hansson et al. (2018) (13) thresholds. In addition to AD-metaROI measures, site was included in the model as a grouping variable for random-effects regression. To assess performance, the area under the receiver operating characteristics curve (AUC) was calculated using the ‘pROC’ package in R. FDR was used to correct for 30 dMRI or 3 cortical thickness tests.

In supplementary analyses, we evaluated classification AUCs of residualized AD-metaROI MRI after adjusting for age, sex, and education. The data were also split into five 80% and 20% train-test subsets and the AUC across the five folds was averaged to assess performance.

#### 2.6.3 Cortical Mediators between CSF Biomarker and Delayed Memory

To better understand the relationship between CSF Aβ_1–42_, pTau_181_, and pTau_181_/Aβ_1–42_ load and cognitive performance, we computed linear regressions with CSF biomarkers as the independent variable and delayed logical memory performance (WMS-R logical memory) as the dependent variable, after adjusting for fixed-effects (age, sex, education) and random-effects (site) covariates. We then completed mediation analyses using the ‘mediation’ package in R to determine whether AD signature AD-metaROI cortical measures (both structural and diffusion) would explain the observed relationships between CSF biomarkers and delayed memory using the same covariate structure (**Supplementary Figure 3**). Per convention, mediation analyses were performed only for AD-metaROI cortical measures that were significantly associated with CSF biomarkers in the primary analyses.

## 3. RESULTS

### 3.1 Participant Demographics

Basic demographic and clinical characteristics of study participants are reported in **Table 1**. There was a significant relationship between continuous CSF biomarker values and clinical diagnosis (Aβ: *F*=3.8 *p*=0.03; pTau: *F*=4.4 *p*=0.02; Aβ/pTau: *F*=9.8 *p*=0.0002), but not sex (Aβ: *F*=3.8 *p*=0.03; pTau: *F*=4.4 *p*=0.02; Aβ/pTau: *F*=9.8 *p*=0.0002). Age was significantly correlated with CSF pTau (*r*=0.39; *p*=0.001) and pTau/AB (*r*=0.30, *p*=0.01), but not Aβ (*r*=-0.17, *p*=0.18), while MMSE was correlated with Aβ (*r*=0.34; *p*=0.006) and pTau/Aβ (*r*=-0.46, *p*=0.0001), but not pTau (*r*=-0.17, *p*=0.18). Education was only correlated with pTau (*r*=-0.26, *p*=0.04), not Aβ (*r*=-0.17, *p*=0.18) or pTau/Aβ (*r*=-0.18, *p*=0.15).

There was no significant relationship between dichotomized CSF Aβ and pTau subgroup (i.e., Aβ-/pTau-, Aβ-/pTau-+, Aβ+/pTau-, Aβ+/pTau+) and clinical diagnosis (*χ*^*2*^=11.6, *p*=0.07), sex (*χ*^*2*^=3.3, *p*=0.35), or age (*F*=0.96, *p*=0.42). However, we found group differences in MMSE (*F*=3.97, *p*=0.01).

### 3.2 Correlations between Cortical AD-metaROI MRI Measures

Pearson’s correlations between cortical AD-metaROI MRI measures revealed that many dMRI measures were highly correlated across participants (**Supplementary Figure 4;** *lower right*). The strongest correlations (Pearson’s *r* > |0.9|) were found between MAP-MRI MSD, QIV, and DTI MD (*r* = 0.92-0.99), and between MAP-MRI RTAP, RTOP, and RTPP (*r* = 0.97-0.99); MAP-MRI RTAP, RTOP, and RTPP were negatively correlated with MAP-MRI MSD and DTI MD (*r* = -0.9--0.97). NODDI ISOVF was also highly correlated with MAP-MRI MSD and DTI MD (*r* = 0.94), and negatively correlated with MAP-MRI RTTP (*r* = -0.94). These dMRI measures remained correlated in the subset of cognitively unimpaired and amyloid negative participants, however CTh was notably less correlated with dMRI measures in this subset (**Supplementary Figure 4;** *top left*).

### 3.3 Primary CSF Biomarker Associations with Regional Cortical MRI Measures

For each CSF and MRI measure, the number of significant cortical ROIs in which a significant association was detected after multiple comparison correction (dMRI *p≤*0.012; CTh *p≤*0.0056) and the direction of associations are reported in **Supplementary Table 1**.

At least one ROI was associated with Aβ_1–42_ across nine out of the 10 dMRI measures evaluated; only ECVF measures were not associated. The largest and most widespread negative associations were detected with QIV (16 ROIs) and ISOVF (15 ROIs) in largely the same temporal and frontal ROIs; MD showed associations in a similar subset of ROIs (13 ROIs; **Figure 1**). The largest effects were detected with QIV in the *pars opercularis, pars triangularis* and insular cortex (*β*=-0.39), followed by the parahippocampal and entorhinal cortex (*β*=-0.38). ICVF was significant in 14 regions but, in addition to frontal and temporal regions, included more parietal cortex associations (largest *β*=0.34 in supramarginal gyrus). The fewest associations were detected with ODI, RTAP, and RTOP; however, one of the largest overall effects was detected with ODI in the supramarginal gyrus (*β*=-0.38).

**Figure 1.**
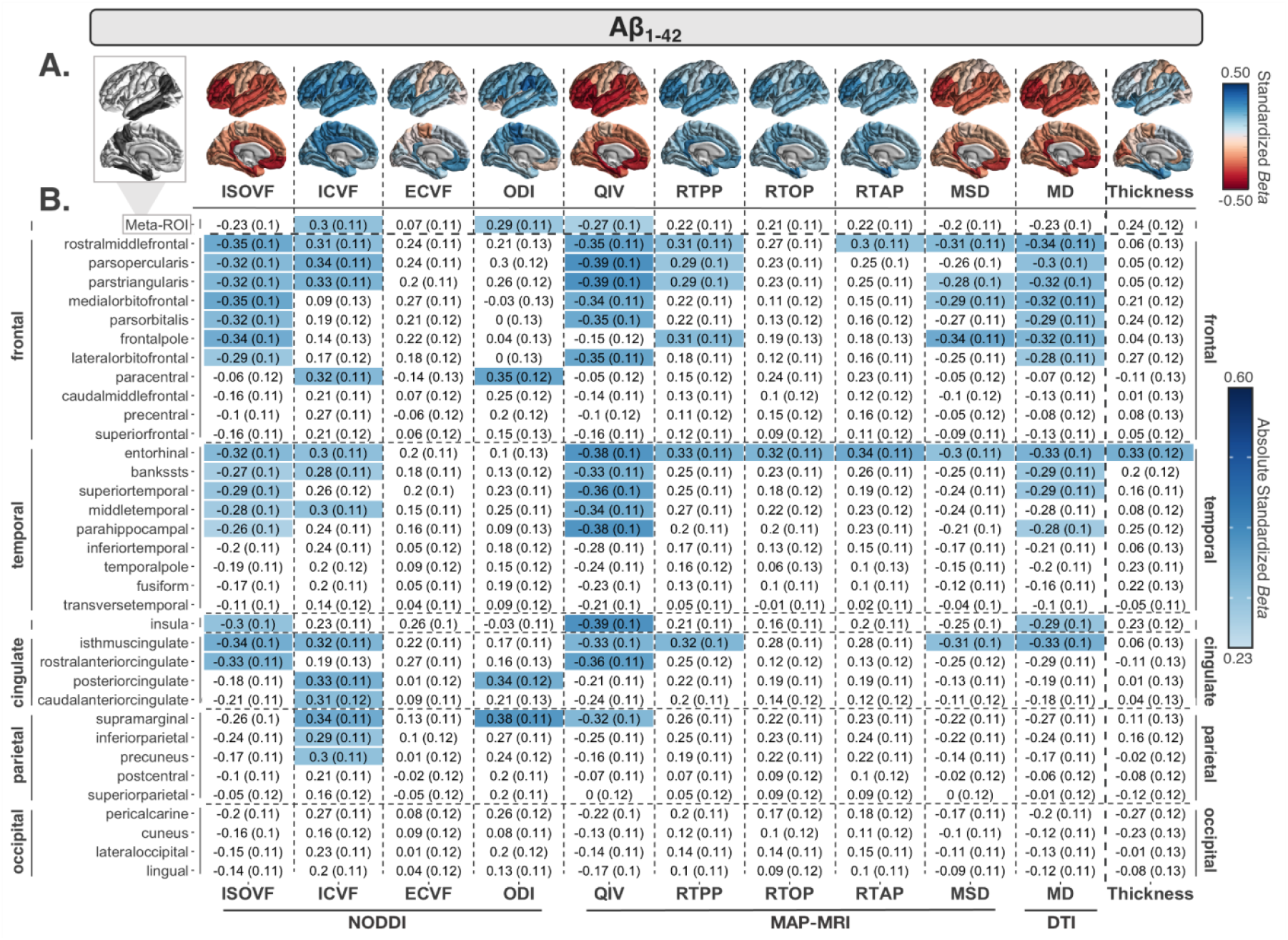
**(A)** Regional effect sizes (*Beta*-values) for associations between CSF Aβ_1–42_ and each cortical measure across participants. For reference, the ROIs comprising the AD-metaROI are highlighted in gray. **(B)** *Beta*-values and standard errors for significant associations (dMRI *p* ≤ 0.012; CTh *p* ≤ 0.0056) are shaded according to the absolute value of their effect size.

Fewer dMRI measures were associated with pTau_181_ than with Aβ_1–42_, but overall effect sizes were larger (**Figure 2**). The largest and most widespread negative associations were detected with ECVF (7 ROIs), followed by positive associations with ISOVF (6 ROIs) and QIV (5 ROIs) in largely frontal and parietal regions. No other dMRI measures were associated with pTau_181_. The greatest overall effects were detected in the paracentral gyrus (ECVF *β*=-0.47; ISOVF *β*=0.40). Similar associations were found when testing log_10_-transformed pTau_181_ (**Supplementary Results 2.3**).

**Figure 2.**
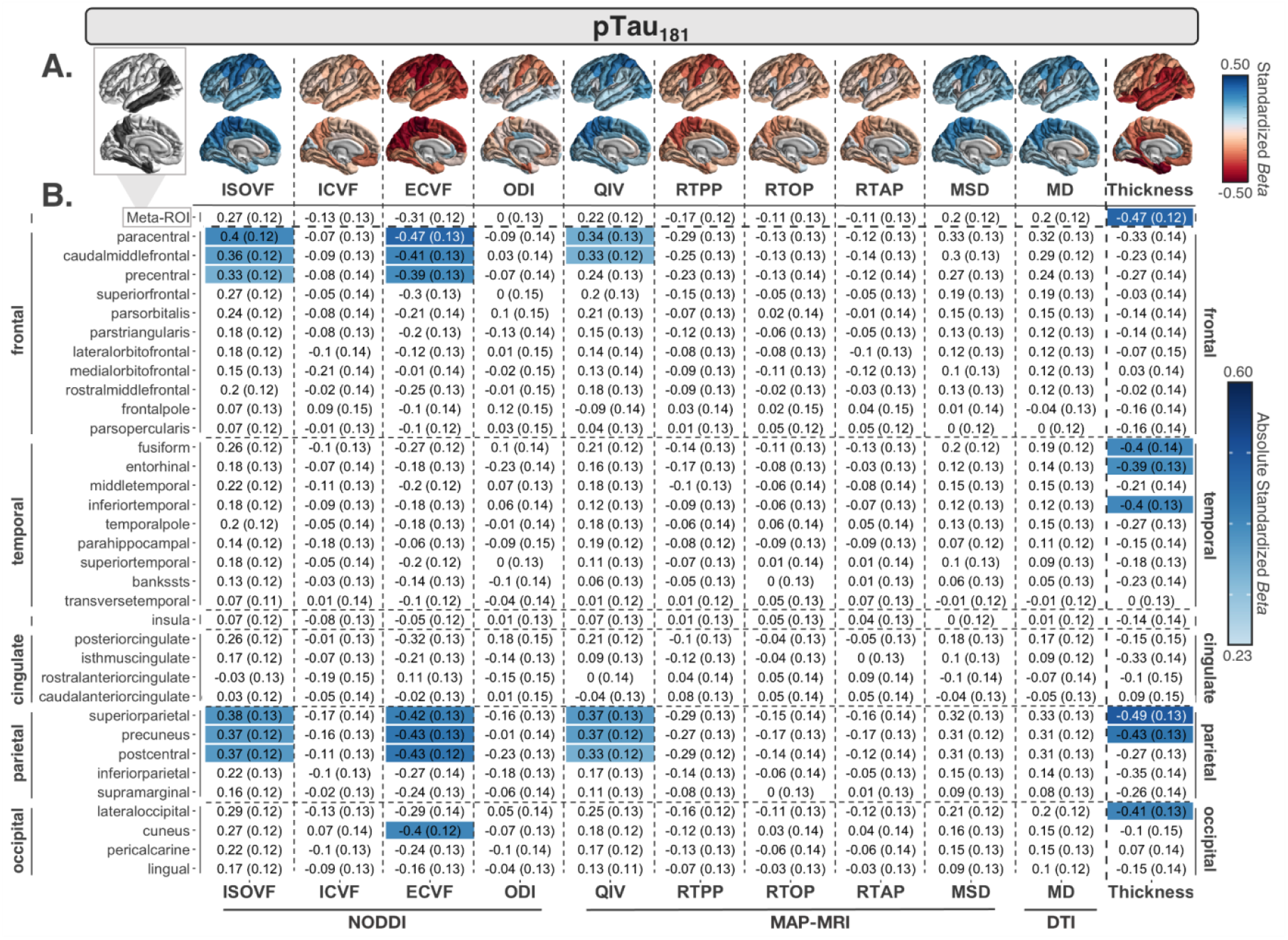
**(A)** Regional effect sizes (*Beta*-values) for associations between CSF pTau_181_ and each cortical measure across participants. For reference, the ROIs comprising the AD-metaROI are highlighted in gray. **(B)** *Beta*-values and standard errors for significant associations (dMRI *p* ≤ 0.012; CTh *p* ≤ 0.0056) are shaded according to the absolute value of their effect size.

More widespread associations were detected with pTau_181_/Aβ_1–42_ across all 10 dMRI measures compared to Aβ_1–42_ or pTau_181_ alone (**Figure 3**). The most widespread positive associations were detected with ISOVF (29 ROIs) and MD (24 ROIs). The largest dMRI effects were detected with QIV in the entorhinal cortex (*β*=0.51), parahippocampal gyrus, and *pars orbitalis* (*β*=0.49). As with Aβ_1–42_, the fewest associations were detected with RTAP, RTOP, ECVF, and ODI.

**Figure 3.**
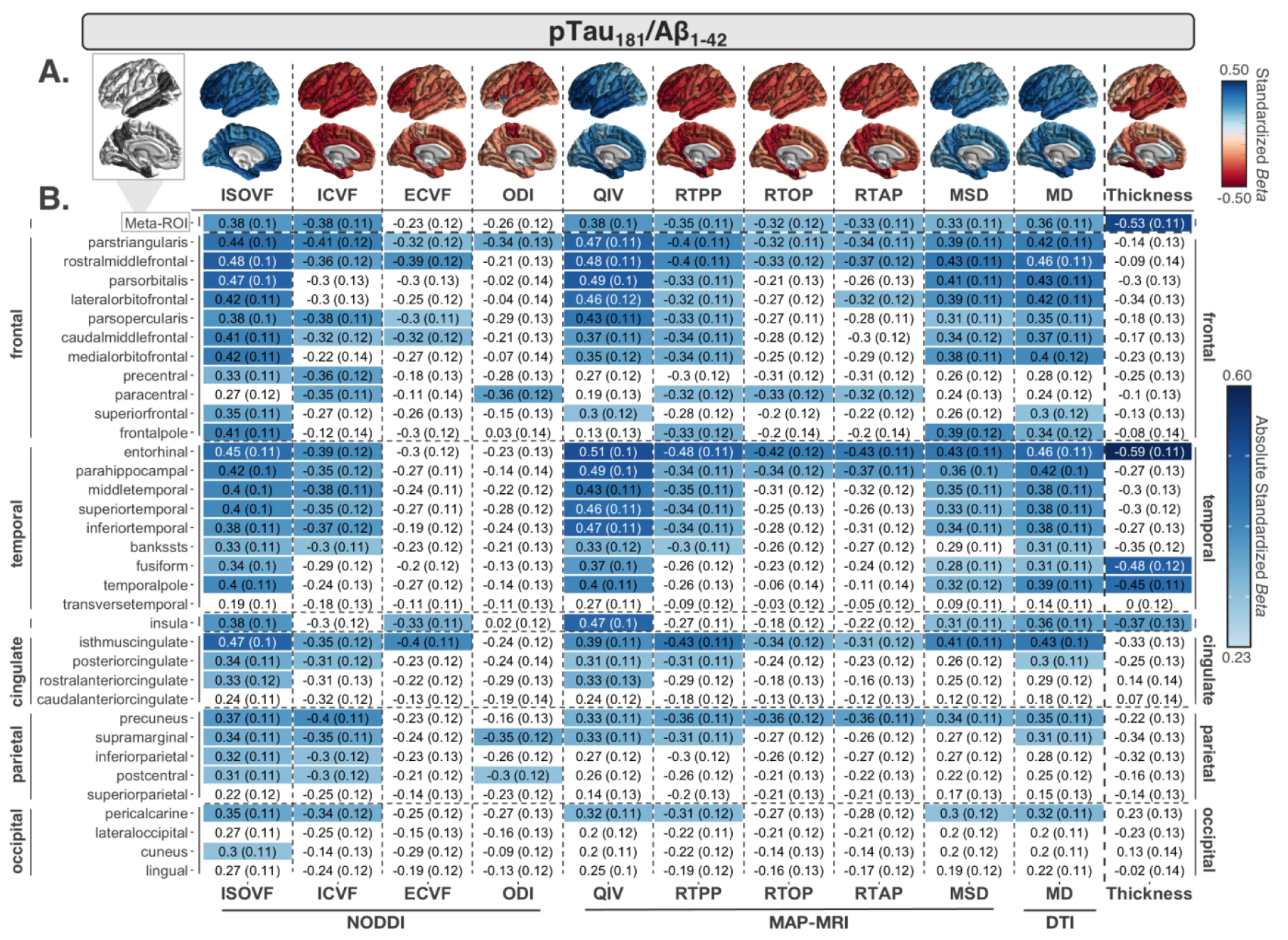
**(A)** Regional effect sizes (*Beta*-values) for associations between CSF pTau_181_/Aβ_1–42_ and each cortical measure across participants. Effect sizes are shown regardless of *P*-value. For reference, the ROIs comprising the AD-metaROI are highlighted in gray. **(B)** *Beta*-values and standard errors for significant associations (dMRI *p* ≤ 0.012; CTh *p* ≤ 0.0056) are shaded according to the absolute value of their effect size.

Significant associations between CTh and both Aβ_1–42_ and pTau_181_/Aβ_1–42_ were more localized compared to the widespread associations detected with cortical dMRI measures; comparable associations were detected with pTau_181_ (7 ROIs). CTh was only associated with Aβ_1–42_ in the entorhinal cortex (*β*=0.33). Across all MRI measures evaluated, however, the largest pTau_181_ and pTau_181_/Aβ_1–42_ associations were detected with CTh in the superior parietal (*β*=-0.49) and entorhinal (*β*=-0.59) cortex, respectively.

In sensitivity analyses, associations between CSF and cortical MRI measures that were significant in the entire cohort were tested in individuals without dementia (i.e., 64 of 66 participants). 98.5% of significant associations remained significant (**Supplementary Results 2.4**).

### 3.4 Secondary AD-metaROI MRI Measure Comparisons between CSF Biomarker Groups

Across all cortical AD-metaROI measures, ANCOVA revealed significant group differences between four dichotomized (+/-) Aβ_1–42_ and pTau_181_ groups (dMRI *p*≤0.041; CTh *p≤*0.0002; **Figure 4 A, B**). In pairwise comparisons, all dMRI measures other than RTOP, RTAP, and ECVF were nominally different between Aβ-/pTau- and Aβ+/pTau+ individuals (*p*<0.05). ISOVF, ICVF, and QIV were nominally different between those Aβ-/pTau+ and Aβ+/pTau+; QIV was also different between Aβ-/pTau- and Aβ+/pTau-individuals. Only CTh differences between Aβ+/pTau+ and the other three groups remained significant after multiple comparisons correction. Overall, there was no detectable difference between pTau- and pTau+; only CTh in Aβ+ individuals was significantly different between pTau- and pTau+.

**Figure 4.**
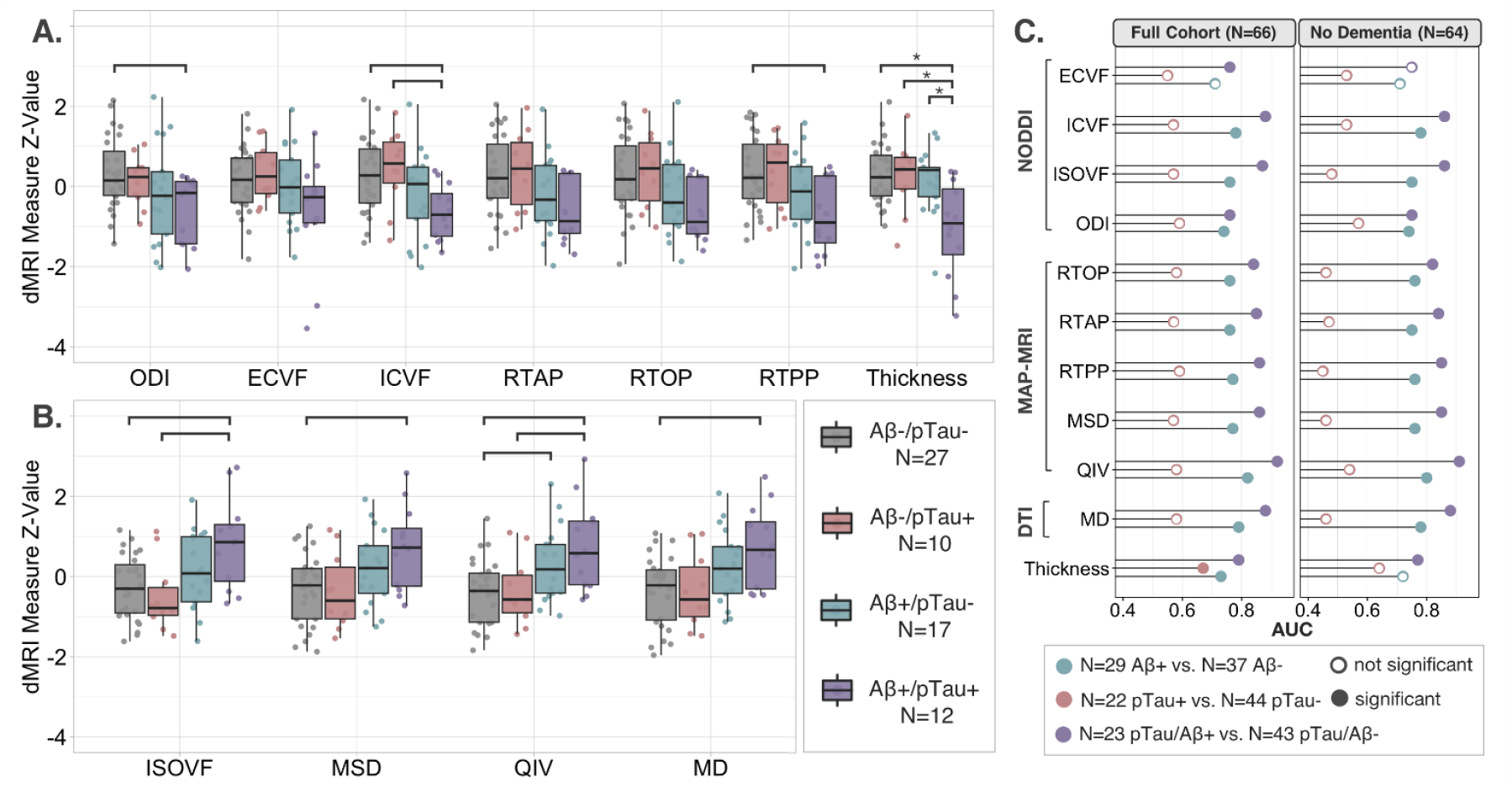
Scaled cortical AD-metaROI measures grouped by CSF biomarker groups show **(A)** lower MT-NODDI ODI, ECVF, ICVF, MAP-MRI RTAP, RTOP, RTPP, and FreeSurfer CTh and **(B)** higher MT-NODDI ISOVF, MAP-MRI MSD, QIV, and DTI MD in Aβ+ compared to Aβ- individuals. The greatest differences are visible between those Aβ-/pTau- and those Aβ+/pTau+; within Aβ- individuals, no major differences are seen between pTau- and pTau+ individuals. Nominal group differences (*p*<0.05) are denoted with brackets; only CTh Aβ+/pTau+ differences were significant after multiple comparisons correction, as denoted with asterisks. **(C)** For each cortical AD-metaROI measure, the area under the curve (AUC) of the dichotomized CSF group classifications are reported. Significant associations after multiple comparisons correction are indicated by filled circles (Full cohort: dMRI *p ≤* 0.021, CTh *p ≤* 0.016; No Dementia: dMRI *p ≤* 0.029, CTh *p ≤* 0.0036)

While underpowered for statistical comparisons (N as low as 2 to 6 participants per group), further breakdown of groups by clinical diagnosis suggests that dMRI effects may be driven by Aβ+ participants with MCI or dementia, regardless of pTau status (**Supplementary Figures 9-10**).

### 3.5 Effect of CSF Biomarker Interactions on AD-metaROI Measures

There were no significant associations between CSF Aβ_1–42_-by-pTau_181_ group (i.e., continuous Aβ_1–42_ by dichotomized pTau_181_) interactions and dMRI AD-metaROI measures. However, a significant Aβ_1–42_-by-pTau_181_ group interaction was found on CTh (*β*=0.64; *p*=0.0051) whereby lower CSF Aβ_1–42_ was associated with lower CTh only in those individuals who were pTau+ (**Supplementary Figure 11**). No significant CSF pTau_181_-by-Aβ_1–42_ group (i.e., continuous pTau_181_ by dichotomized Aβ_1–42_) interactions were detected for any AD-metaROI measure (**Supplementary Figure 12**).

### 3.6 Exploratory Classification of CSF Biomarker Groups by Cortical MRI Measures

As reported in **Figure 4B** and **Supplementary Table 4**, except for ECVF, dMRI AD-metaROI measures better distinguished Aβ+ from Aβ-participants than did CTh (dMRI AUC=0.74-0.82; CTh AUC=0.73); QIV performed best (AUC=0.82) while MD, ICVF, RTPP, and MSD performed within 0.05 of QIV (AUC=0.77-0.79). In contrast, only CTh significantly classified the pTau_181_ groupings (AUC=0.67). While all MRI measures were significant, pTau_181_/Aβ_1–42_ group was also best classified by QIV (AUC=0.92) with comparable performance (i.e., within 0.05) by ICVF, ISOVF and MD (AUC=0.87-0.88). Similar classification trends were found 1) in the subset of participants without dementia, 2) when AD-metaROI MRI measures were first residualized (i.e., adjusted for age, sex, and education), and 3) with five-fold cross-validation using 80/20 training/test splits (**Supplementary Results 2.7**).

### 3.7 Cortical MRI Mediators between CSF Biomarker and Delayed Memory

Lower CSF Aβ_1–42_ was associated with worse delayed memory performance (*p*=0.01; *β*=0.30), as hypothesized. In previous analyses, Aβ_1–42_ was significantly associated with QIV, ICVF, and ODI AD-metaROI measures (**Figure 1**). The effect of Aβ_1–42_ on memory was fully mediated by ICVF (33% of the total effect) and QIV (41%; **Table 2**).

**Table 2.** AD-metaROI cortical mediators between CSF markers and delayed memory. Only AD-metaROI measures associated with Aβ_1–42_ or pTau_181_/Aβ_1–42_ in primary analyses were evaluated; we corrected for multiple comparisons across 12 mediation tests. The *direct effect* reflects the effect of CSF on delayed memory. The *mediation effect* reflects the effect of CSF on memory that goes through the cortical mediator. Effect sizes (*β*), uncorrected *p*-values, and the percent of the total CSF effect on memory (direct + indirect) that was mediated are reported.

| CSF | MRI Model | Cortical Mediator | Direct Effect |  | Mediation Effect |  |  |
| --- | --- | --- | --- | --- | --- | --- | --- |
| | | | $\beta$ | P | $\beta$ | P | % |
| **$A\beta$**<br>$p=0.01$<br>$\beta=0.30$ | MT-NODDI | ICVF | 0.21 | 0.078 | 0.10 | **0.014\*\*** | 32.7 |
|  |  | ODI | 0.28 | **0.026\*\*** | 0.02 | 0.592 | 6.4 |
|  | MAP-MRI | QIV | 0.17 | 0.14 | 0.12 | **0.008\*\*** | 41.2 |
| **pTau/$A\beta$**<br>$p=5.9 \times 10^{-5}$<br>$\beta=-0.50$ | MT-NODDI | ICVF | -0.40 | **<2x10<sup>-16</sup>\*\*** | -0.10 | **0.036\*\*** | 19.8 |
|  |  | ISOVF | -0.36 | **0.004\*\*** | -0.14 | **0.008\*\*** | 27.6 |
|  | MAP-MRI | MSD | -0.38 | **<2x10<sup>-16</sup>\*\*** | -0.12 | **0.012\*\*** | 22.1 |
|  |  | QIV | -0.34 | **0.006\*\*** | -0.13 | **0.01\*\*** | 27.2 |
|  |  | RTAP | -0.40 | **<2x10<sup>-16</sup>\*\*** | -0.10 | **0.014\*\*** | 20.2 |
|  |  | RTOP | -0.42 | **<2x10<sup>-16</sup>\*\*** | -0.09 | **0.026\*\*** | 17.4 |
|  |  | RTPP | -0.39 | **<2x10<sup>-16</sup>\*\*** | -0.11 | **0.02\*\*** | 22.2 |
|  | DTI | MD | -0.36 | **0.004\*\*** | -0.12 | **0.01\*\*** | 25.8 |
|  | FreeSurfer | CTh | -0.37 | **0.006\*\*** | -0.11 | 0.12 | 23.4 |

\*\* $p < \text{FDR Threshold for 12 tests}$ ; Direct $p \leq 0.026$ ; Mediation $p \leq 0.036$

Greater pTau_181_/Aβ_1–42_ was also associated with worse delayed memory performance (*p*=5.9×10^−5^; *β* =-0.50). CSF pTau_181_/Aβ_1–42_ was significantly associated with CTh and all AD-metaROI measures except ODI and ECVF (**Figure 3**). Each of the seven dMRI AD-metaROI measures was found to partially mediate the effect of pTau_181_/Aβ_1–42_ on delayed memory (∼17-28% of the total effect). CTh did not have a mediating effect (**Table 2**).

We did not find significant associations between CSF pTau_181_ or log_10_-transformed pTau_181_ and memory. Mediation results from sensitivity analyses excluding individuals with dementia (N=2) revealed similar results to those in the whole sample (**Supplementary Table 8**).

## 4. DISCUSSION

In this study, we investigated the relationship between regional microstructural dMRI measures in cortical gray matter and CSF Aβ and pTau in a population of largely cognitively unimpaired and MCI individuals from the third phase of the Alzheimer’s Disease Neuroimaging Initiative. We found that lower CSF Aβ and higher CSF pTau were associated with cortical DTI, NODDI and MAP-MRI dMRI measures reflecting less hindered or restricted diffusion (i.e., lower RTAP, RTOP, RTPP, ICVF) and greater diffusivity (i.e., higher MD, MSD, QIV, ECVF, ISOVF). Cortical dMRI measures were more widely associated with Aβ than pTau and better distinguished Aβ+ from Aβ-participants. In contrast, cortical thickness was more tightly linked with pTau. dMRI associations mediated the relationship between CSF markers and performance on logical memory tasks, which is vulnerable to AD pathogenesis.

### 4.1 CSF Aβ Associations with Cortical MRI Measures

Cortical dMRI measures were widely associated with CSF amyloid concentrations, particularly in AD vulnerable regions such as the entorhinal, isthmus cingulate, and frontal cortex. These extensive associations are consistent with patterns of both amyloid PET tracer uptake (45-47) and post-mortem amyloid deposits found throughout the neocortex (48) in early to intermediate disease stages.

Across all dMRI measures, QIV from MAP-MRI showed the most robust associations with CSF Aβ and it best distinguished Aβ+ from Aβ-participants. QIV is a general measure of the variance of the diffusion signal, which has been shown to be highly sensitive to the presence of biological barriers in regions with lower restriction and greater isotropic volume fractions values, such as in the cortical ribbon (35); conversely, RTOP and RTAP (also derived from MAP-MRI) may only be sensitive in regions with extremely high restriction such as the WM. MAP-MRI is fitted to the overall diffusion signal (i.e, from both intra- and extra-cellular compartments) and imposes few assumptions, potentially offering better sensitivity to amyloid load than multi-compartment models such as NODDI.

NODDI parses the signal into intra- and extra-cellular spaces and resulting ICVF, ECVF, and ISOVF measures may be sensitive to different aspects of AD histopathology. Measures reflecting lower diffusion restriction (i.e., ICVF) may reflect loss of cortical neurites, while lower ODI (also derived from NODDI) may be associated with lower complexity in dendritic arborization. Greater ISOVF or free-water volume may be a function of both neurodegeneration and greater inflammation-associated edema (49-51). Inflammation plays a substantial role in AD pathogenesis (52). Persistent immune response in the brain is not only associated with neurodegeneration but exacerbates Aβ and tau pathology and may even link initial Aβ pathology to subsequent NFT development (53).

ICVF and ISOVF also showed spatially distinct patterns of associations with amyloid; while ICVF associations were strongest in fronto-temporal regions, ISOVF associations also included parietal regions. Among regions that were selectively associated with each measure, the largest effect size differences between ICVF and ISOVF amyloid associations were found in either the earliest or latest myelinating regions of the neocortex: ISOVF showed larger effects in medial orbitofrontal cortex (late/lightly myelinated), while ICVF showed larger effects in paracentral cortex (early/heavily myelinated; **Supplementary Figure 16**). These measures may capture the proposed inverse relationship between the pattern of pathology progression and myelination during development (54, 55). Selective associations between ISOVF and amyloid in late myelinating regions that are vulnerable to early pathogenesis may reflect more advanced degeneration in these regions, however, it is also possible that compared to ICVF, ISOVF is a better suited metric for regions that simply lack sufficient myelin. NODDI may fail to properly estimate measures of restriction in unmyelinated tissue (56).

Although amyloid plaques are formed in the extracellular space, we found no associations between CSF Aβ and ECVF. This may be partially due to the dMRI acquisition parameters used in ADNI, where the diffusion time is t_d_=30.5 ms and the maximum b-value is b=2000 s/mm^2^. When dMRI sequences do not have sufficiently short diffusion times (t_d_ ≤ 20 ms) and high b-values (b>3000 s/mm^2^), the NODDI model is unable to differentiate the extracellular space from permeable cell bodies due to water exchange. Sequences with such advanced parameters can be used to fit models such as cell body and neurite density imaging (SANDI) (57), which can also estimate cell body size and density. These sequences can currently only be acquired on one of the few existing ultra-high gradient strength Connectome scanners (57, 58), and are not currently feasible for large-scale multisite clinical studies of neurodegenerative disorders.

Our results, however, support the use of measures derived from common single-shell sequences, such as DTI MD, for associations with CSF amyloid. We found both MD regional associations and classification performance were comparable to multi-shell ISOVF. Higher MD and ISOVF measures may be particularly confounded by exacerbated partial volume effects in the presence of cortical atrophy, so we mitigated these effects by excluding voxels with ISOVF>0.5 from all mean cortical dMRI measures (59). In fact, AD-metaROI MD and ISOVF were more tightly correlated with ICVF than cortical thickness, suggesting they capture microstructural phenomenon beyond the partial voluming that can be attributable to atrophy.

In contrast to dMRI measures, cortical thickness was only associated with amyloid in the entorhinal cortex. Aβ biomarker abnormalities are thought to precede tau and neurodegeneration in many proposed models of the AD pathological cascade. T1w macrostructural measures, such as cortical thickness, are the most frequently used biomarkers of neurodegeneration, however our results support evidence that changes in microstructural dMRI measures can precede those detectable with T1w MRI and may therefore be better suited for detecting early cortical Aβ pathology.

### 4.2 CSF pTau Associations with Cortical MRI Measures

Compared to amyloid, diffusion measures showed fewer regional associations with CSF pTau, albeit larger effect sizes. These effects were restricted to frontal and parietal regions implicated in later *Braak stages, 5-6*. This aligns with our preliminary comparisons between CSF biomarker groups which show that pTau effects appear to be driven only by Aβ+ individuals (N=12), particularly those with MCI or dementia (N=6; **Supplementary Figures 9-10**), suggesting a later disease stage. These preliminary comparisons are highly limited by the number of individuals in each group, which may explain the limited number of regional pTau associations detected in our study.

Counterintuitively, given the evidence showing *intracellular* pTau accumulation that results in cell body NFTs, the most robust dMRI effects detected were lower ECVF. Although the specific mechanisms are unknown, accumulation and aggregation of pTau in the intracellular space (i.e., cell body), in conjunction with extracellular pathology (e.g., amyloid plaques, inflammatory cytokines, ghost tangles) may together create enough barriers to hinder diffusion captured by ECVF. Since cell death occurs heterogeneously in the cortex, the lack of ICVF associations may be driven by opposing effects of intracellular pTau aggregation in some cells increasing restriction (i.e, increased ICVF) and the death of other cells decreasing restriction (i.e, decreasing ICVF). The observed association between higher ISOVF with greater pTau in late Braak regions could be the result of a greater degree of neuronal loss in later stages. As one of the first studies examining these effects, the mechanisms driving our findings are still unknown; however, increases in both cortical ICVF and ISOVF have been previously reported in studies of human tauopathy mouse models (rTg4510) (60).

Higher pTau was most strongly associated with lower cortical thickness in the superior parietal cortex and metaROI. Early *Braak 1-4* temporal and metaROI pTau associations were only detected with cortical thickness, perhaps suggesting more advanced atrophy in regions with earlier involvement. Medial temporal tau increases with age and does not necessarily indicate AD pathology (61). In contrast, significant pTau associations in the latest *Braak 6* regions, including postcentral, paracentral, cuneus, and precentral cortex, were only detected with microstructural dMRI measures, which may capture earlier stages of degeneration. Larger studies will allow for event based modeling to estimate the sequence in which these cortical biomarkers become abnormal (62). This approach can be used to help build an understanding of the dynamics of not only typical disease progression, but also to identify distinct spatiotemporal trajectories of tau subtypes (63).

### 4.3 CSF pTau/Aβ Associations with Cortical MRI Measures

As expected, we found the largest overall associations between cortical MRI measures and CSF pTau/Aβ. Cortical measures also better classified dichotomized CSF ratios than individual Aβ or pTau measures. While most of the associations between imaging and CSF ratios aligned with results when examining Aβ and pTau separately, MAP-MRI measure associations were more widespread than those observed using the measures individually, consistent with studies showing better diagnostic accuracy of CSF biomarkers when analyzed together (64, 65).

### 4.4 Cortical MRI Mediators between CSF Biomarker and Delayed Memory

While both CSF Aβ and pTau/Aβ were associated with delayed memory, pTau/Aβ had larger effects. This supports the large body of literature showing that amyloid burden does not correlate well with clinical severity and that both amyloid and tau are necessary for cognitive decline (66, 67). We found that dMRI measures mediated a larger portion of Aβ effects (up to 41%) than pTau/Aβ (up to 28%), in line with the limited direct relationship between amyloid and cognitive outcomes compared to tau. Amyloid may drive downstream cognitive impairment through microstructural damage. Interestingly, only dMRI measures mediated the relationship between pTau/Aβ and memory while cortical thickness did not. Given that the sample was largely cognitively unimpaired, this aligns with our findings that dMRI measures are more sensitive to earlier pathological changes such as amyloid deposition.

There is a large body of literature showing that pTau is more tightly linked with neurodegeneration and clinical severity than Aβ, however we did not find significant associations between CSF pTau and memory. This may be due in part to the limited number of participants in our study, particularly those with high pTau concentrations. As shown in **Supplementary Figure 17** two CU individuals (one Aβ+ and one Aβ-) with high delayed memory scores and high pTau were sufficient to drive the association (or lack thereof).

### 4.5 Comparison to other studies

Several AD dMRI studies of preclinical asymptomatic individuals have suggested a non-monotonic or biphasic relationship between AD pathology and dMRI measures of brain microstructure (Weston et al. 2015), including two studies specifically evaluating 1) associations between CSF amyloid and tau biomarkers and either single-shell free-water and DTI MD (11) or 2) NODDI ICVF and ODI cortical measures (8). An early transient stage of increased diffusion restriction or lower diffusivity followed by a later phase of lower restriction and increased diffusivity has been identified in dMRI studies of both GM (68) and WM (69, 70). These studies propose that lower diffusivity associations with greater amyloid burden, measured with CSF or PET, in the earliest stages could be attributable to factors such as cellular hypertrophy or inflammatory microglia infiltration and proliferation increasing the number of diffusion barriers. Subsequent associations showing higher diffusivity with greater amyloid load may be driven by cellular loss and neurodegeneration. Our findings in a mix of CU and MCI participants are in line with the second part of this trajectory. Future work evaluating a larger sample of CU participants is needed to isolate earlier signatures. As CSF changes may occur before changes in PET are detectable, using both data types would further allow for the examination of the earliest diffusion trajectories in CSF+/PET-participants.

### 4.6. Limitations and Future Directions

There are several limitations to this study. First, the small number of ADNI participants with multi-shell dMRI data limited our ability to parse specific neuroimaging patterns within each diagnostic and biomarker group. Second, the ADNI3 dMRI acquisition does not support the standard NODDI modeling assumptions in the GM, therefore, additional studies are also needed across heterogeneous dMRI acquisitions (i.e., higher b-values/shorter diffusion times) to replicate these findings. Our results may be considered preliminary until replicated in additional larger, independent cohorts. Third, while our mediation models were based on theoretical models of causality, we cannot infer a causal relationship given the cross-sectional dataset. Future work will investigate this relationship using a longitudinal design. Finally, our “AD signature” metaROI was based on cortical thickness associations with Braak neurofibrillary tangle stage at autopsy (44), potentially biasing our classification and mediation analyses. There is a need to define meta-ROIs based on associations between dMRI measures and amyloid.

### 4.7 Conclusions

Understanding the relationship between various dMRI measures of brain microstructure and Aβ and tau pathology may improve upon current AD models and provide further insight into mechanisms underlying cognitive decline. Diffusion MRI metrics, calculated across multiple models, may better track amyloid progression through non-invasive means, compared to cortical thickness, the most commonly used measure of cortical damage.

## Data Availability

The study used openly available human data from the Alzheimer's Disease Neuroimaging Initiative (ADNI) database (https://ida.loni.usc.edu/)

## ACKNOWLEDGMENTS

This work was supported by the National Institutes of Health (grant numbers T32AG058507, P41EB015922, R01AG058854, R01AG059874, RF1AG057892). It was also funded in part by a U.S. Alzheimer’s Association Zenith Award.

Data collection and sharing for this project was funded by the Alzheimer’s Disease Neuroimaging Initiative (ADNI) (National Institutes of Health Grant U01 AG024904) and DOD ADNI (Department of Defense award number W81XWH-12-2-0012). ADNI is funded by the National Institute on Aging, the National Institute of Biomedical Imaging and Bioengineering, and through generous contributions from the following: AbbVie, Alzheimer’s Association; Alzheimer’s Drug Discovery Foundation; Araclon Biotech; BioClinica, Inc.; Biogen; Bristol-Myers Squibb Company; CereSpir, Inc.; Cogstate; Eisai Inc.; Elan Pharmaceuticals, Inc.; Eli Lilly and Company; EuroImmun; F. Hoffmann-La Roche Ltd and its affiliated company Genentech, Inc.; Fujirebio; GE Healthcare; IXICO Ltd.;Janssen Alzheimer Immunotherapy Research & Development, LLC.; Johnson & Johnson Pharmaceutical Research & Development LLC.; Lumosity; Lundbeck; Merck & Co., Inc.;Meso Scale Diagnostics, LLC.; NeuroRx Research; Neurotrack Technologies; Novartis Pharmaceuticals Corporation; Pfizer Inc.; Piramal Imaging; Servier; Takeda Pharmaceutical Company; and Transition Therapeutics. The Canadian Institutes of Health Research is providing funds to support ADNI clinical sites in Canada. Private sector contributions are facilitated by the Foundation for the National Institutes of Health (www.fnih.org). The grantee organization is the Northern California Institute for Research and Education, and the study is coordinated by the Alzheimer’s Therapeutic Research Institute at the University of Southern California. ADNI data are disseminated by the Laboratory for NeuroImaging at the University of Southern California.

## Conflict of Interest Statement

MWW has served on the scientific advisory boards for Lilly, Araclon, and Institut Catala de Neurociencies Aplicades, Gulf War Veterans Illnesses Advisory Committee, VACO, Biogen Idec, and Pfizer; has served as a consultant for Astra Zeneca, Araclon, Medivation/Pfizer, Ipsen, TauRx Therapeutics Ltd., Bayer Healthcare, Biogen Idec, Exonhit Therapeutics, SA, Servier, Synarc, Pfizer, and Janssen; has received funding for travel from NeuroVigil, Inc., CHRU-Hopital Roger Salengro, Siemens, AstraZeneca, Geneva University Hospitals, Lilly, University of California, San Diego–ADNI, Paris University, Institut Catala de Neurociencies Aplicades, University of New Mexico School of Medicine, Ipsen, CTAD (Clinical Trials on AD), Pfizer, AD PD meeting, Paul Sabatier University, Novartis, Tohoku University; has served on the editorial advisory boards for Alzheimer’s and Dementia and MRI; has received honoraria from NeuroVigil, Inc., Insitut Catala de Neurociencies Aplicades, PMDA/Japanese Ministry of Health, Labour, and Welfare, and Tohoku University; has received commercial research support from Merck and Avid; has received government research support from DOD and VA; has stock options in Synarc and Elan; and declares the following organizations as contributors to the Foundation for NIH and thus to the NIA funded AD Neuroimaging Initiative Abbott, Alzheimer’s Association, Alzheimer’s Drug Discovery Foundation, Anonymous Foundation, AstraZeneca, Bayer Healthcare, BioClinica, Inc. (ADNI 2), Bristol-Myers Squibb, Cure Alzheimer’s Fund, Eisai, Elan, Gene Network Sciences, Genentech, GE Healthcare, GlaxoSmithKline, Innogenetics, Johnson and Johnson, Eli Lilly and Company, Medpace, Merck, Novartis, Pfizer Inc., Roche, Schering Plough, Synarc, and Wyeth.

CRJ has provided consulting services for Janssen Research & Development, LLC., and Eli Lilly. MB is a former employee of GE Medical Systems and receives pension payment. CJ consults for Lily and serves on an independent data monitoring board for Roche but he receives no personal compensation from any commercial entity. CJ receives research support from NIH and the Alexander Family Alzheimer’s Disease Research Professorship of the Mayo Clinic.

PMT and NJ report a research grant from Biogen, Inc. for research unrelated to this manuscript.

The remaining authors declare that the research was conducted in the absence of any commercial or financial relationships that could be construed as a potential conflict of interest.

## Notes

### Author Declarations

The study used ONLY openly available human data from the Alzheimer's Disease Neuroimaging Initiative (ADNI) database (https://ida.loni.usc.edu/)

