## Supplementary Appendix for "Cortical microstructural associations with CSF amyloid and pTau"

Supplementary Table 3
Supplementary Figure 6
Supplementary Figure 7
Supplementary Figure 8
Supplementary Table 4
Supplementary Figure 13
Supplementary Table 5
Supplementary Figure 14
Supplementary Table 6
Supplementary Figure 15
Supplementary Table 7
Supplementary Figure 17

### 1. Supplementary Methods

#### 1.1 Cortical dMRI measure extraction flow chart

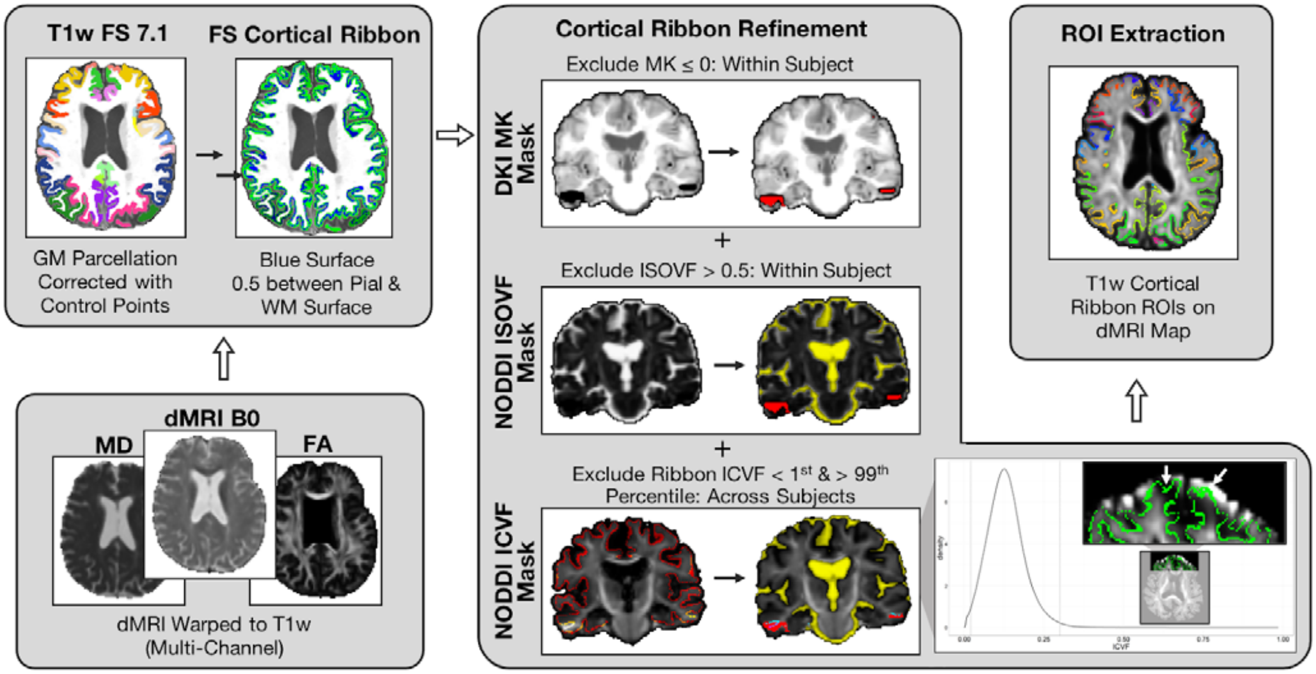

**Supplementary Figure 1.** After spatially normalizing dMRI maps to respective parcellated T1w images, mean dMRI measures were extracted from a cortical ribbon halfway between the pial and white matter surfaces. To further mitigate any remaining artifacts, partial volume effects, and misregistration the following were excluded from the cortical ribbon: (1) voxels with implausible negative kurtosis values, i.e., DKI MK value ≤ 0; (2) voxels with CSF partial voluming, i.e., ISOVF > 0.5; and (3) as illustrated, voxels in the ribbon that had extreme ICVF values for the sample, i.e., voxels with ICVF values in the 1$^{st}$ and 99$^{th}$ percentiles across all participants. These tended to occur in regions with uncorrected EPI induced distortions such as the frontal and temporal lobes.

##### 1.2 MT-NODDI parallel diffusivity MSE comparisons

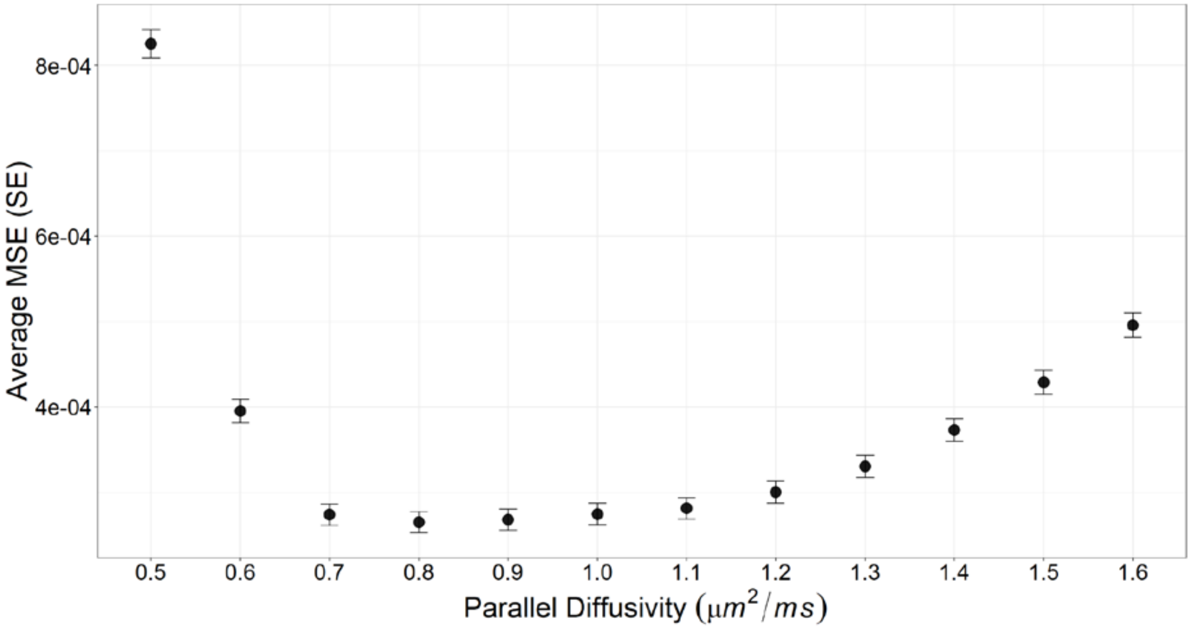

**Supplementary Figure 2.** The average mean square error (MSE), for each $d_{\parallel}$, between MT-NODDI measured and predicted signals averaged within the medial cortical ribbon across CU participants. The lowest error was found with $d_{\parallel} = 0.8$ μ$m^2$/ms. Paired two-sided t-tests revealed cortical MSE for $d_{\parallel} = 0.8$ was significantly lower than all other $d_{\parallel}$ values.

##### 1.3 Proposed mediation analysis

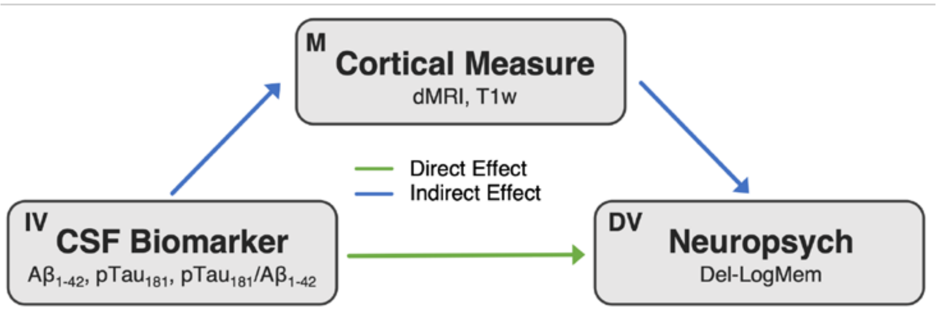

**Supplementary Figure 3.** Proposed mediation analysis. *Key: IV: Independent Variable; M: Mediator; DV: Dependent Variable.*

#### 2. Supplementary Results

##### 2.1 Correlation between cortical AD-metaROI measures

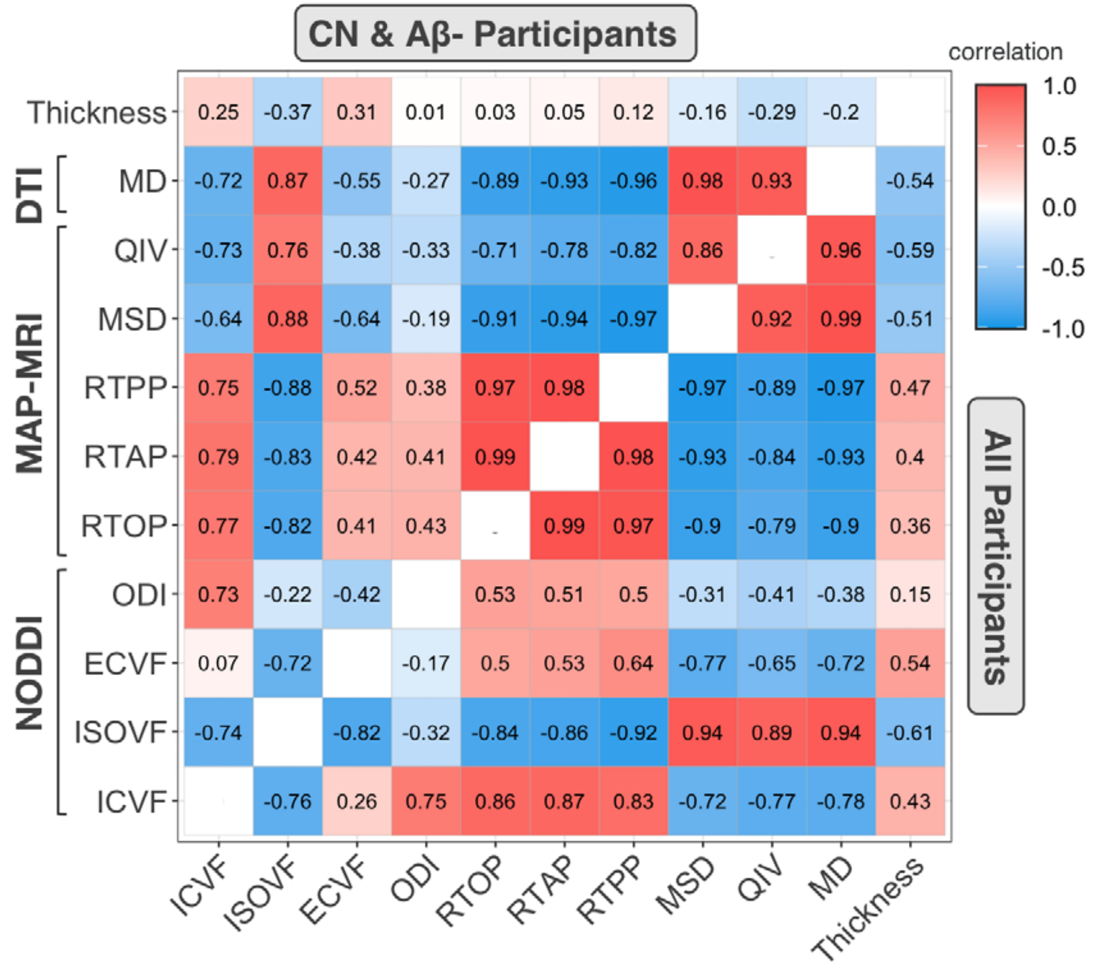

**Supplementary Figure 4.** Pearson's correlation coefficients between cortical AD-metaROI measures across all participants (N=66; *lower right*) and across the subset of $A\beta_{1-42}$ negative and CU participants (N=29; *upper left*).

#### 2.2 CSF biomarker associations with cortical MRI measures

**Supplementary Table 1.** Number of significant cortical ROIs-- out of 34 individual ROIs and the AD signature meta-ROI tested -- and direction of associations between each cortical MRI and CSF measure (dMRI $p \leq 0.012$; CTh $p \leq 0.0056$).

| Model | Measure | Aβ | | pTau | | pTau/Aβ | | Total N |
|---|---|---|---|---|---|---|---|---|
| | | N | +/- | N | +/- | N | +/- | |
| MT-NODDI | ECVF | | | 7 | - | 6 | - | 13 |
| | ISOVF | 15 | - | 6 | + | 29 | + | 50 |
| | ICVF | 14 | + | | | 20 | - | 34 |
| | ODI | 4 | + | | | 4 | - | 8 |
| MAP-MRI | MSD | 6 | - | | | 20 | + | 26 |
| | QIV | 16 | - | 5 | + | 24 | + | 45 |
| | RTAP | 2 | + | | | 9 | - | 11 |
| | RTOP | 1 | + | | | 8 | - | 9 |
| | RTPP | 6 | + | | | 21 | - | 27 |
| DTI | MD | 13 | - | | | 24 | + | 37 |
| FreeSurfer | CTh | 1 | + | 7 | - | 5 | - | 13 |
| Total N | | 78 | | 25 | | 170 | | 273 |

#### 2.3 Log10-transformed pTau181 associations

**Supplementary Table 2.** Summary of log10 transform pTau results. Number of significant cortical ROIs-- out of 34 individual ROIs and the AD signature meta-ROI tested -- and direction of associations between each cortical MRI and CSF measure. When using the log10 transform of pTau instead of pTau, the threshold for significant dMRI associations did not change (dMRI $p \leq 0.012$), while CTh did (CTh $p \leq 0.00086$). Compared to a total of 273 significant associations when assessing Aβ, pTau, and pTau/Aβ, a total of 268 significant associations were detected across Aβ, $\log_{10}$(pTau), and pTau/Aβ.

| Model | Measure | Aβ | | $\log_{10}$(pTau) | | pTau/Aβ | | Total N |
|---|---|---|---|---|---|---|---|---|
| | | N | +/- | N | +/- | N | +/- | |
| MT-NODDI | ECVF | | | 9 | - | 6 | - | 15 |
| | ISOVF | 15 | - | 6 | + | 29 | + | 50 |
| | ICVF | 14 | + | | | 20 | - | 34 |
| | ODI | 4 | + | | | 4 | - | 8 |
| MAP-MRI | MSD | 6 | - | 2 | + | 20 | + | 28 |
| | QIV | 16 | - | 1 | + | 24 | + | 41 |
| | RTAP | 2 | + | | | 9 | - | 11 |
| | RTOP | 1 | + | | | 8 | - | 9 |
| | RTPP | 6 | + | | | 21 | - | 27 |
| DTI | MD | 13 | - | 1 | + | 24 | + | 38 |
| FreeSurfer | CTh | | | 3 | - | 4 | - | 7 |
| Total N | | 77 | | 22 | | 169 | | 268 |

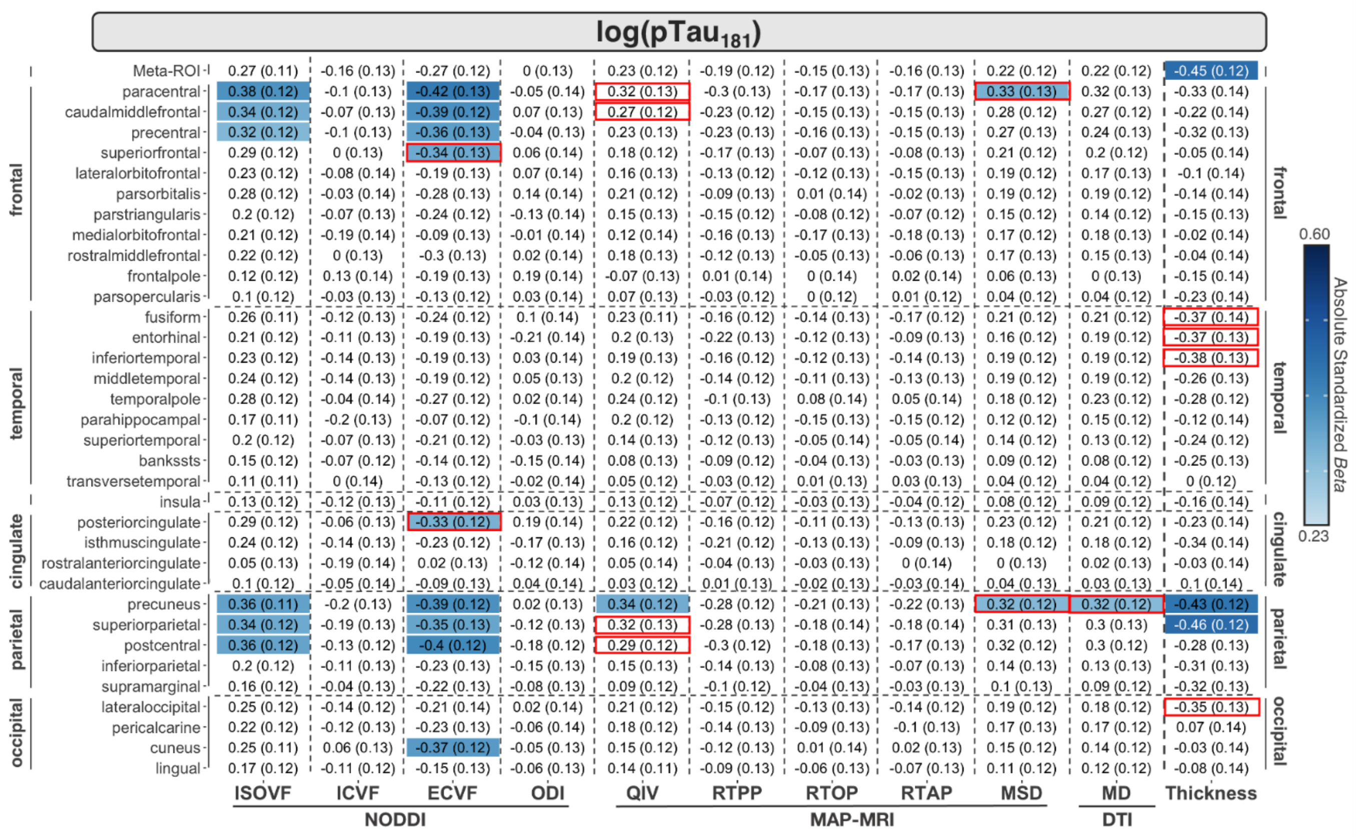

**Supplementary Figure 5**. Effect sizes (*Beta*-values and standard error) for associations between log10-transformed CSF $pTau_{181}$ and each cortical measure. Significant associations (dMRI $p \leq 0.012$; CTh $p \leq 0.00086$) are shaded according to the absolute value of their effect size. For comparison with $pTau_{181}$, ROIs that changed significance are highlighted in red boxes.

#### 2.4 Sensitivity analysis: CSF biomarker associations in non-demented participants

**Supplementary Table 3.** Number of significant ROIs (dMRI $p \leq 0.048$; CTh $p \leq 0.029$) and direction of associations between each cortical and CSF measure in the subset of participants without dementia (N=64). Only measures and ROIs that were significant in the entire cohort were evaluated. The percent of the ROIs significant in the non-demented subset relative to the whole cohort is reported. Only four of 170 p-tau$_{181}$/Aβ$_{1–42}$ ROIs that were significant in the full cohort were not significant in non-demented participants.

| Model | Measure | Aβ$_{1–42}$ | | | p-tau$_{181}$ | | | p-tau$_{181}$/Aβ$_{1–42}$ | | | Total N (%) |
|---|---|---|---|---|---|---|---|---|---|---|---|
| | | N | % | +/- | N | % | +/- | N | % | +/- | |
| MT-NODDI | ECVF | | | | 7 | 100 | - | 6 | 100 | - | 13 (100%) |
| | ISOVF | 15 | 100 | - | 6 | 100 | + | 29 | 100 | - | 50 (100%) |
| | ICVF | 14 | 100 | + | | | | 20 | 100 | - | 34 (100%) |
| | ODI | 4 | 100 | + | | | | 4 | 100 | - | 8 (100%) |
| MAPMRI | MSD | 6 | 100 | - | | | | 20 | 100 | + | 26 (100%) |
| | QIV | 16 | 100 | - | 5 | 100 | + | 22 | 91.7 | + | 43 (95.6%) |
| | RTAP | 2 | 100 | + | | | | 9 | 100 | - | 11 (100%) |
| | RTOP | 1 | 100 | + | | | | 8 | 100 | - | 9 (100%) |
| | RTPP | 6 | 100 | + | | | | 20 | 95.2 | - | 26 (96.3%) |
| DTI | MD | 13 | 100 | - | | | | 23 | 95.8 | + | 36 (97.3%) |
| FreeSurfer | CTh | 1 | 100 | + | 7 | 100 | - | 5 | 100 | - | 13 (100%) |
| Total N (%) | | 78 (100%) | | | 25 (100%) | | | 166 (97.6%) | | | 269 (98.5%) |

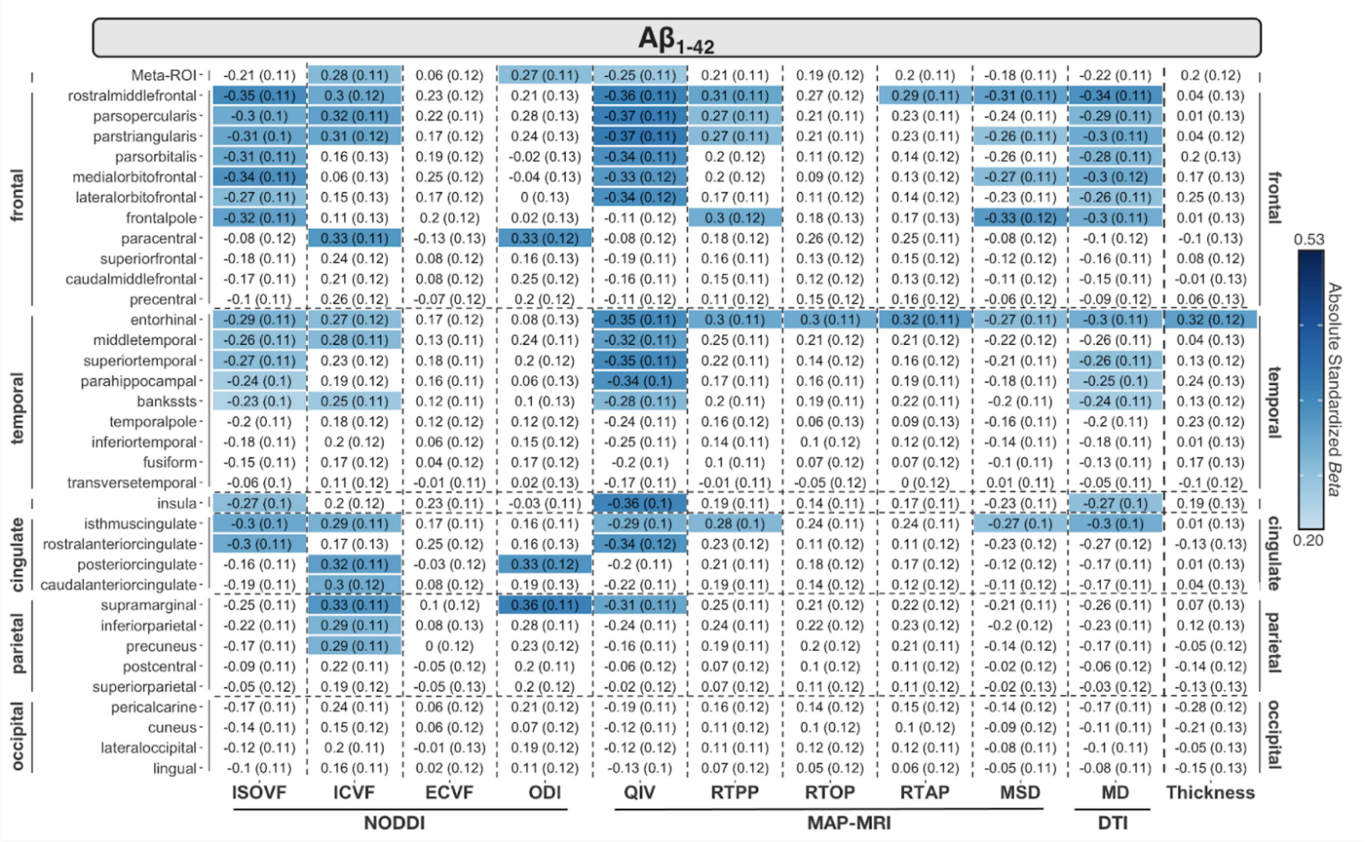

**Supplementary Figure 6**. Effect sizes (*Beta*-values and standard error) for associations between CSF $A\beta_{1-42}$ and each cortical measure in the subset of participants without dementia (N=64). Significant associations (dMRI $p \leq 0.048$; CTh $p \leq 0.029$) are shaded according to the absolute value of their effect size; only measures and ROIs that were significant in the entire cohort were considered. All ROIs that were significant in the full cohort remained significant in non-demented participants.

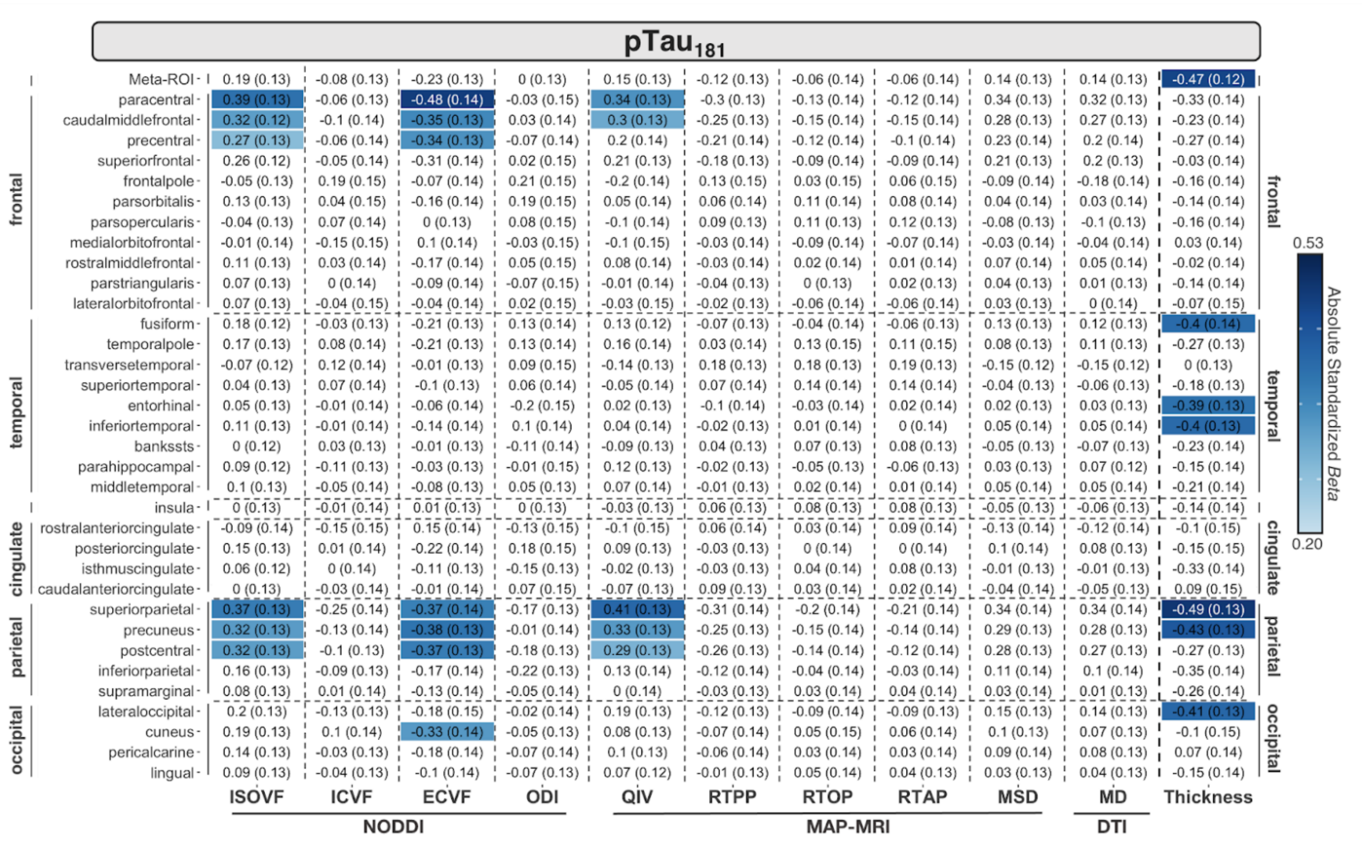

**Supplementary Figure 7**. Effect sizes (*Beta*-values and standard error) for associations between CSF pTau$_{181}$ and each cortical measure in the subset of participants without dementia (N=64). Significant associations (dMRI $p \leq 0.048$; CTh $p \leq 0.029$) are shaded according to the absolute value of their effect size; only measures and ROIs that were significant in the entire cohort were considered. All ROIs that were significant in the full cohort remained significant in non-demented participants.

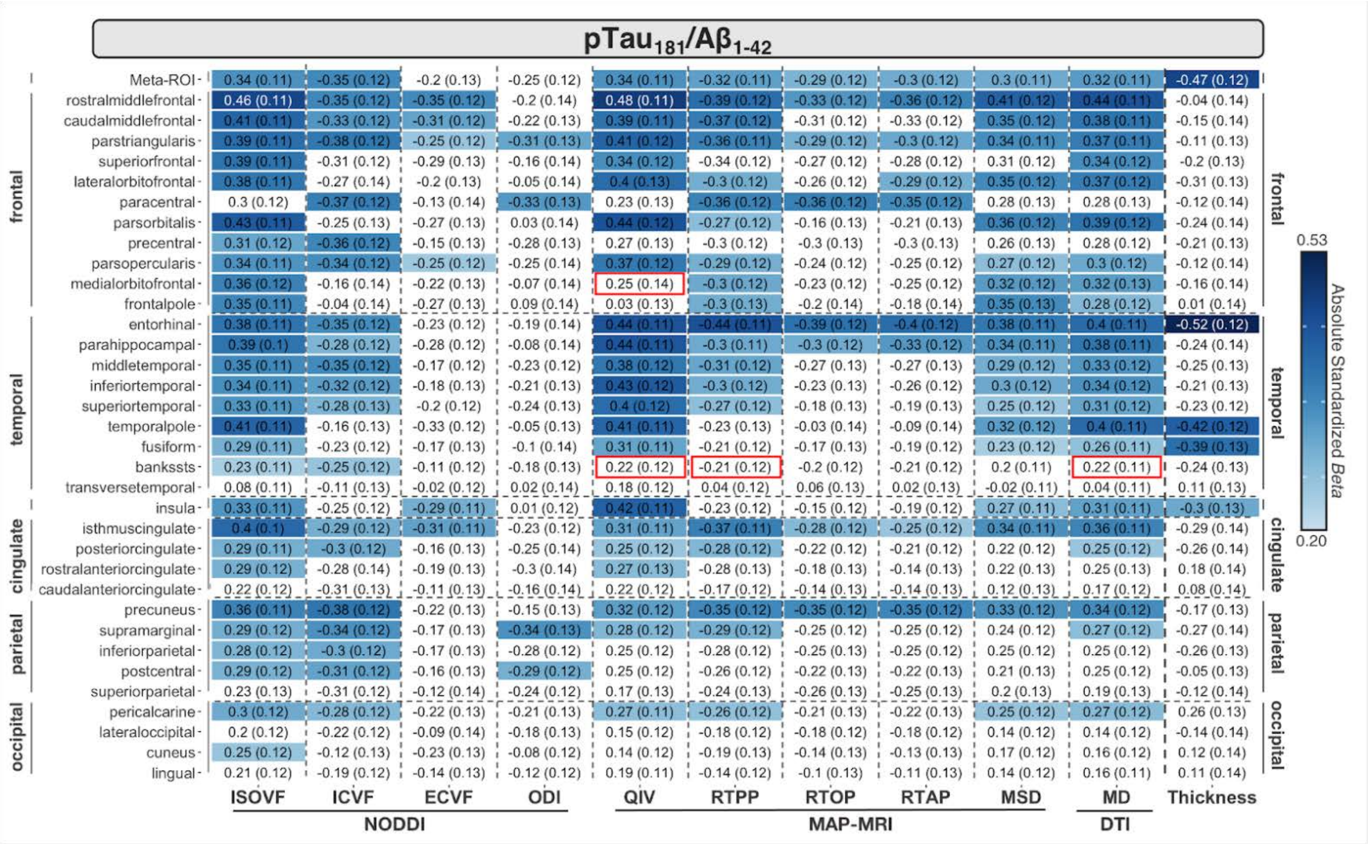

**Supplementary Figure 8**. Effect sizes (*Beta*-values and standard error) for associations between CSF $pTau_{181}/A\beta_{1-42}$ and each cortical measure in the subset of participants without dementia (N=64). Significant associations (dMRI $p \leq 0.048$; CTh $p \leq 0.029$) are shaded according to the absolute value of their effect size; only measures and ROIs that were significant in the entire cohort were considered. Only four ROIs that were significant in the full cohort were not significant in non-demented participants; these are highlighted in red boxes.

#### 2.5 AD-metaROI MRI differences between CSF and clinical diagnosis group

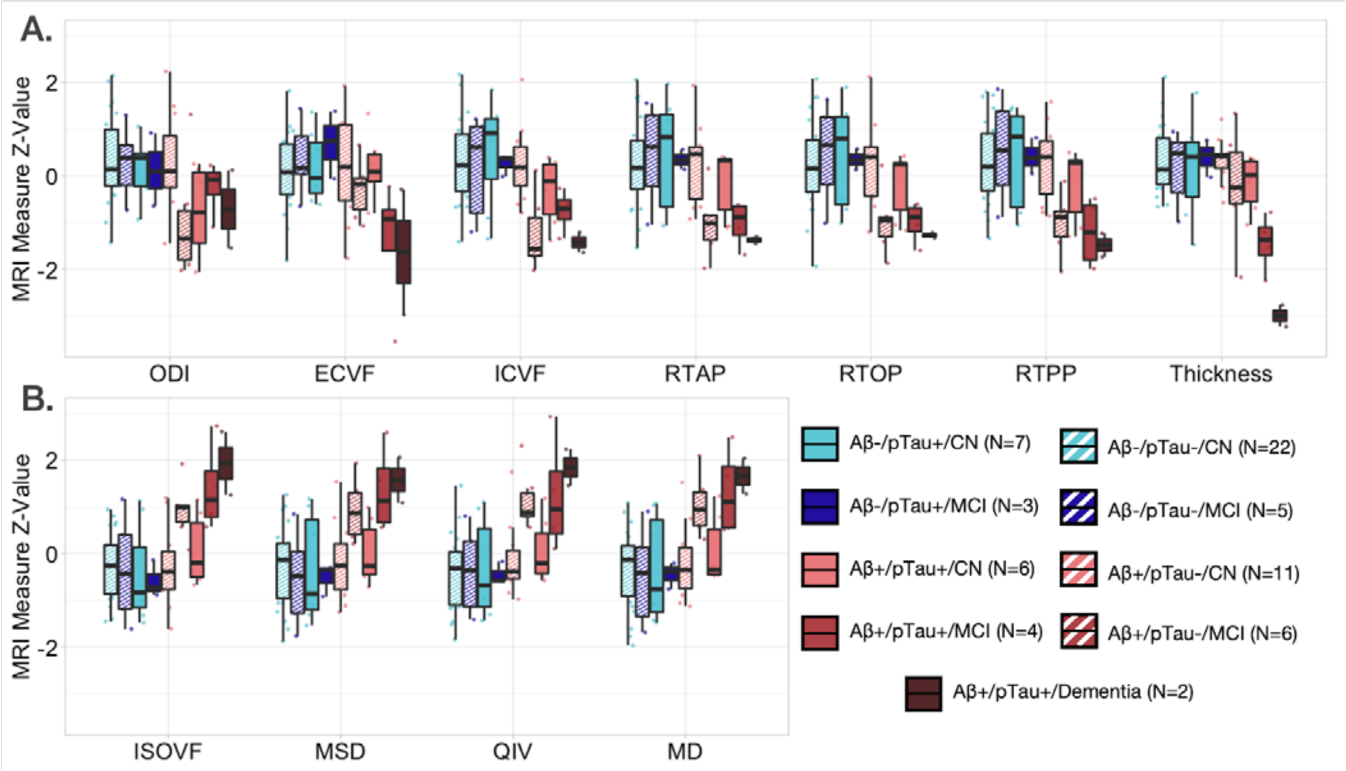

**Supplementary Figure 9**. Scaled cortical AD-metaROI MRI measures classified based on four CSF biomarker cutoff subgroups and three clinical diagnoses show **(A)** lower MT-NODDI ODI, ECVF, ICVF, MAPMRI RTAP, RTOP, RTPP, and FreeSurfer CTh (*top panel*) and **(B)** higher MT-NODDI ISOVF, MAPMRI MSD, QIV, and DTI MD (*bottom panel*) in Aβ+ (*reds*) compared to Aβ- (*blues*) individuals. While group sizes are very small (as low as 2 to 6 participants), the greatest metaROI differences are visible in Aβ+ participants with either MCI or Dementia, regardless of pTau status (pTau- depicted in *stripes* while pTau+ are *solid*).

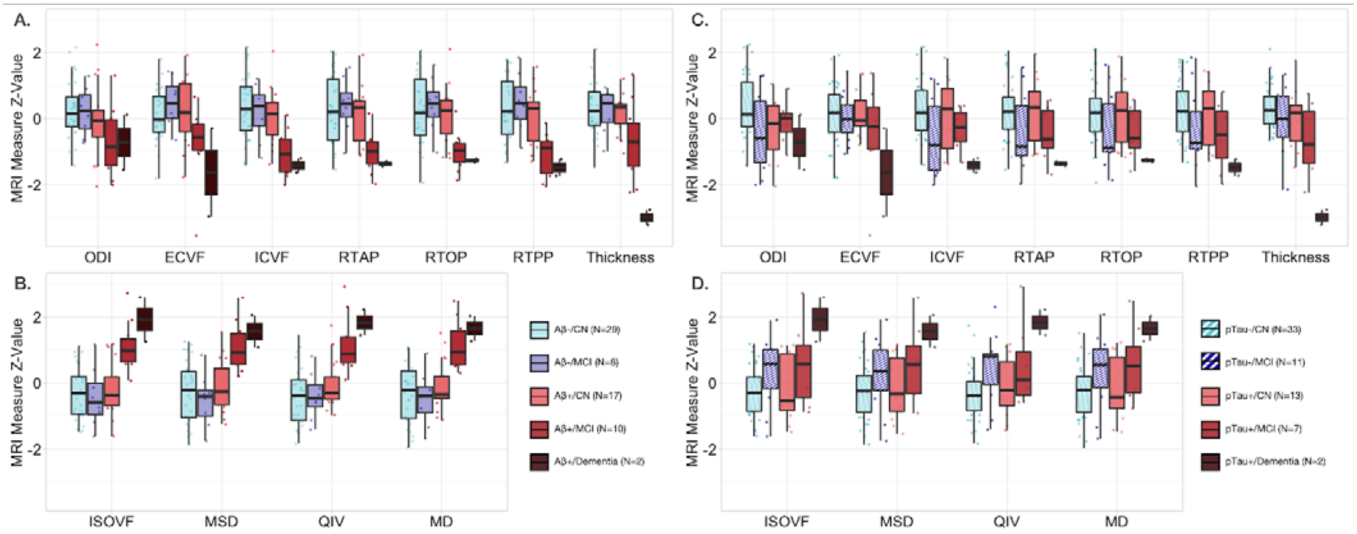

**Supplementary Figure 10**. Scaled cortical AD-metaROI measures classified based on **(A)** Aβ+/- cutoff subgroups (*reds* vs *blues*) show the greatest dMRI differences in Aβ+ participants with either MCI or dementia (*left*). **(B)** Scaled metaROI measures classified based on pTau +/- cutoff subgroups (*solid* vs *striped*) only show large dMRI differences in the two pTau+ participants with dementia, while MCI participants show moderate differences regardless of pTau status *(right)*.

#### 2.6 Interactive effects of CSF biomarker on AD-metaROI MRI measures

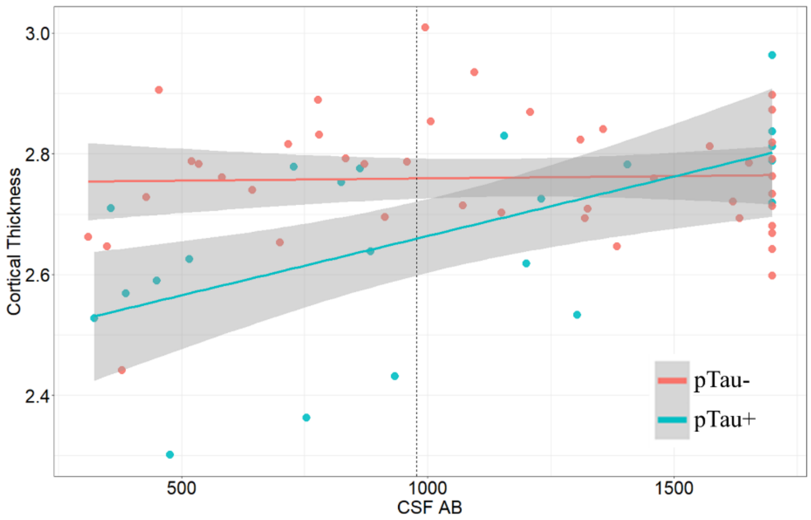

**Supplementary Figure 11.** The relationship between AD-metaROI cortical thickness and CSF Aβ concentration was significantly moderated by pTau group (pTau+/-). The dotted line on the x-axis indicates the CSF Aβ cut point for positivity.

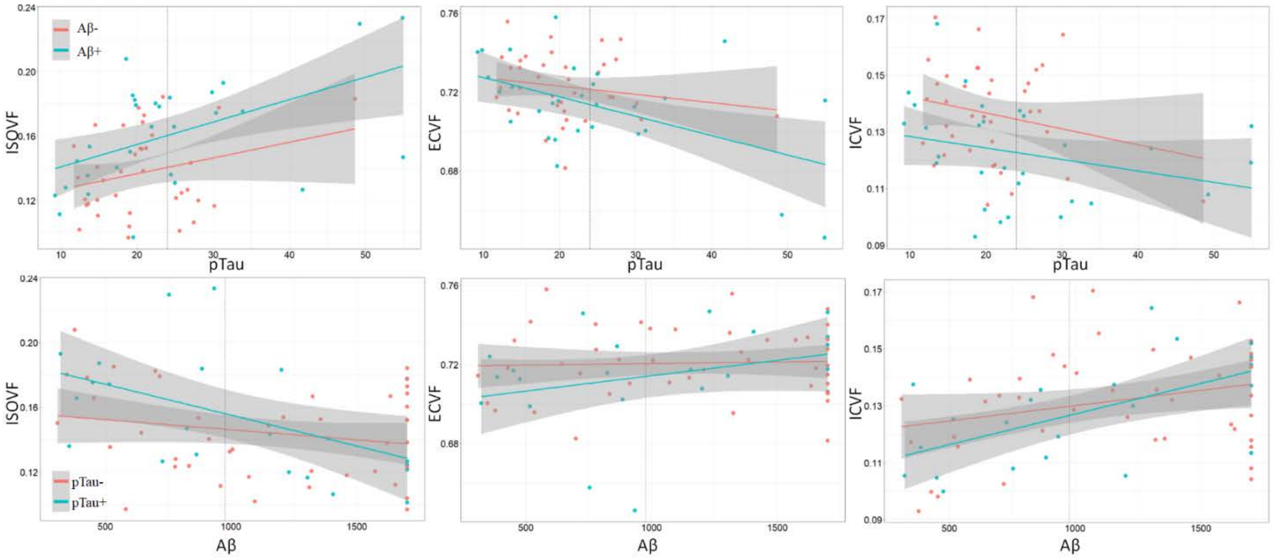

**Supplementary Figure 12**. While no significant interactive effects were detected, the relationship between AD-metaROI NODDI measures and CSF pTau concentration in either Aβ+ or Aβ- participants (*top row*) or CSF Aβ concentration in either pTau+ or pTau- participants (*bottom row*) are shown for reference. Dotted lines on the x-axis indicate the CSF biomarker cut points for each measure.

#### 2.7 Exploratory CSF biomarker group classification by cortical AD-metaROI measures

##### 2.7.1 Non-residualized cortical AD-metaROI measure CSF group classification

###### 2.7.1.1 Logistic regression classification across participants

**Supplementary Table 4.** For each cortical AD-metaROI measure (not residualized), dichotomized CSF biomarker classification area under the curve (AUC)

| Model | Measure | Full Cohort (N=66) | | | Exclude Dementia (N=64) | | |
|---|---|---|---|---|---|---|---|
| | | Aβ | pTau | pTau/Aβ | Aβ | pTau | pTau/Aβ |
| MT-NODDI | ECVF | 0.71 | 0.55 | 0.76** | 0.71 | 0.53 | 0.75* |
| | ICVF | 0.78** | 0.57 | 0.88** | 0.78** | 0.53 | 0.86** |
| | ISOVF | 0.76** | 0.57 | 0.87** | 0.75** | 0.48 | 0.86** |
| | ODI | 0.74** | 0.59 | 0.76** | 0.74** | 0.57 | 0.75** |
| MAP-MRI | RTOP | 0.76** | 0.58 | 0.84** | 0.76** | 0.46 | 0.82** |
| | RTAP | 0.76** | 0.57 | 0.85** | 0.75** | 0.47 | 0.84** |
| | RTPP | 0.77** | 0.59 | 0.86** | 0.76** | 0.45 | 0.85** |
| | MSD | 0.77** | 0.57 | 0.86** | 0.76** | 0.46 | 0.85** |
| | QIV | **0.82**** | 0.58 | **0.92**** | **0.80**** | 0.54 | **0.91**** |
| DTI | MD | 0.79** | 0.58 | 0.88** | 0.78** | 0.46 | 0.88** |
| FreeSurfer | Thickness | 0.73** | **0.67**** | 0.79** | 0.72* | **0.64** | 0.77** |

**Full cohort: dMRI $p \leq 0.021$, CTh $p \leq 0.016$
**No Dementia: dMRI $p \leq 0.029$, CTh $p \leq 0.0036$
*p<0.05

###### 2.7.1.2 Five-fold logistic regression classification 80-20 split

As reported in **Supplementary Figure 13** and **Supplementary Table 5**, dMRI AD-metaROI measures better distinguished Aβ+ from Aβ- participants than did cortical thickness (mean AUC=0.66); ISOVF, RTPP, MSD, and MD performed similarly (AUC=0.75-0.77) and QIV performed marginally better (AUC=0.79). In contrast, for $pTau_{181}$ classification, cortical thickness (AUC=0.69) and dMRI ISOVF, RTOP, RTAP, RTPP, QIV and MD (AUC=0.65-0.66) performed marginally better than ECVF, ICVF, and ODI. $pTau_{181}/A\beta_{1-42}$ status was also better classified by dMRI measures (AUC=0.87-0.89) than cortical thickness (AUC=0.76), excluding ECVF and ODI. Similar classification trends were found in the subset of participants without dementia, the most notable differences being that compared to the full group 1) measures of restriction and ODI better classified Aβ status, and 2) there was an even more notable difference in $pTau_{181}$ classification accuracy between diffusion and thickness measures, with diffusion measures performing on average marginally worse than in the full group and cortical thickness performing better (AUC=0.76).

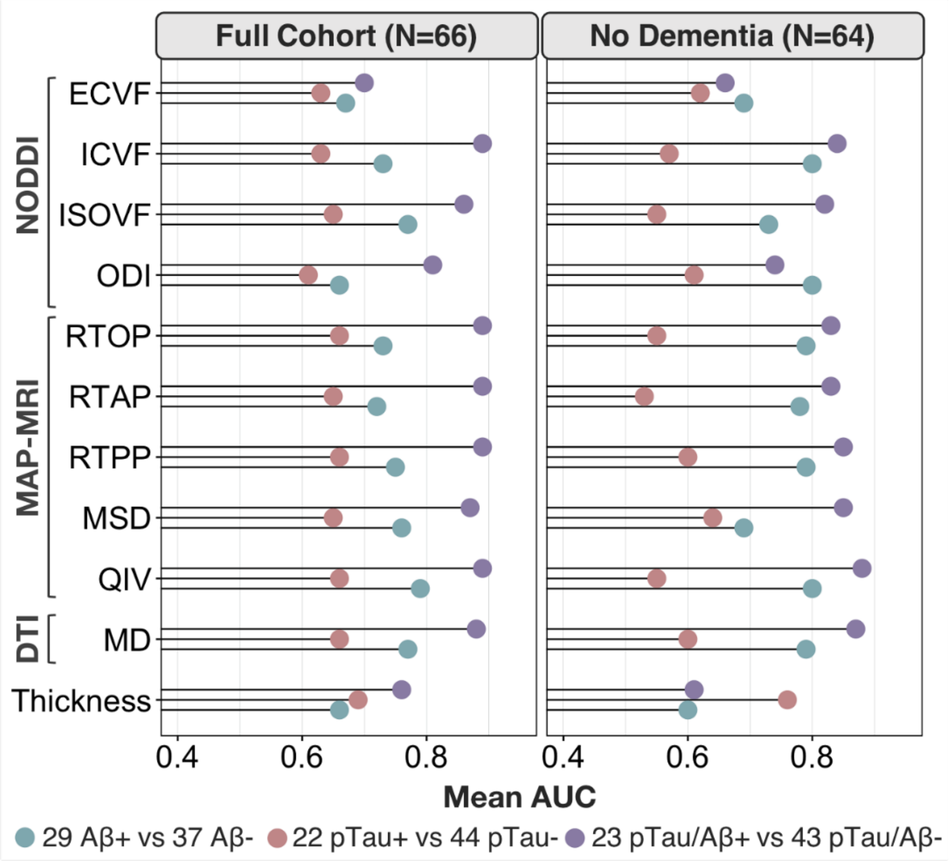

**Supplementary Figure 13.** For each cortical AD-metaROI measure, the average dichotomized CSF biomarker group classification area under the curve (AUC) from 5 folds is reported.

**Supplementary Table 5.** For each cortical AD-metaROI measure (not residualized), the average dichotomized CSF biomarker classification area under the curve (AUC) from 5 folds is reported.

| Model | Measure | Full Cohort (N=66) | | | Exclude Dementia (N=64) | | |
|---|---|---|---|---|---|---|---|
| | | **Aβ** | **pTau** | **pTau/Aβ** | **Aβ** | **pTau** | **pTau/Aβ** |
| **MT-NODDI** | **ECVF** | 0.67 | 0.63 | 0.70 | 0.69 | 0.62 | 0.66 |
| | **ICVF** | 0.73 | 0.63 | **0.89** | **0.80** | 0.57 | 0.84 |
| | **ISOVF** | 0.77 | 0.65 | 0.86 | 0.73 | 0.55 | 0.82 |
| | **ODI** | 0.66 | 0.61 | 0.81 | **0.80** | 0.61 | 0.74 |
| **MAP-MRI** | **RTOP** | 0.73 | 0.66 | **0.89** | 0.79 | 0.55 | 0.83 |
| | **RTAP** | 0.72 | 0.65 | **0.89** | 0.78 | 0.53 | 0.83 |
| | **RTPP** | 0.75 | 0.66 | **0.89** | 0.79 | 0.60 | 0.85 |
| | **MSD** | 0.76 | 0.65 | 0.87 | 0.69 | 0.64 | 0.85 |
| | **QIV** | **0.79** | 0.66 | **0.89** | **0.80** | 0.55 | **0.88** |
| **DTI** | **MD** | 0.77 | 0.66 | 0.88 | 0.79 | 0.60 | 0.87 |
| **FreeSurfer** | **CTh** | 0.66 | **0.69** | 0.76 | 0.60 | **0.76** | 0.61 |

##### 2.7.2 Residualized cortical AD-metaROI measure CSF group classification

###### 2.7.2.1 Logistic regression classification across participants

As reported in **Supplementary Figure 14** and **Supplementary Table 6**, residualized dMRI AD-metaROI measures (i.e., adjusted for age, sex, and education significantly distinguished Aβ+ from Aβ- participants with similar performance from ICVF, ISOVF, RTPP, MSD, and MD (AUC=0.77-0.79), and QIV performing marginally better (AUC=0.82). In contrast, only cortical thickness significantly classified $pTau_{181}$ (AUC=0.70). $pTau_{181}/A\beta_{1-42}$ status was also best classified by ICVF (AUC=0.81), MD (AUC=0.82), and QIV (AUC=0.85). Similar classification trends were found in the subset of participants without dementia.

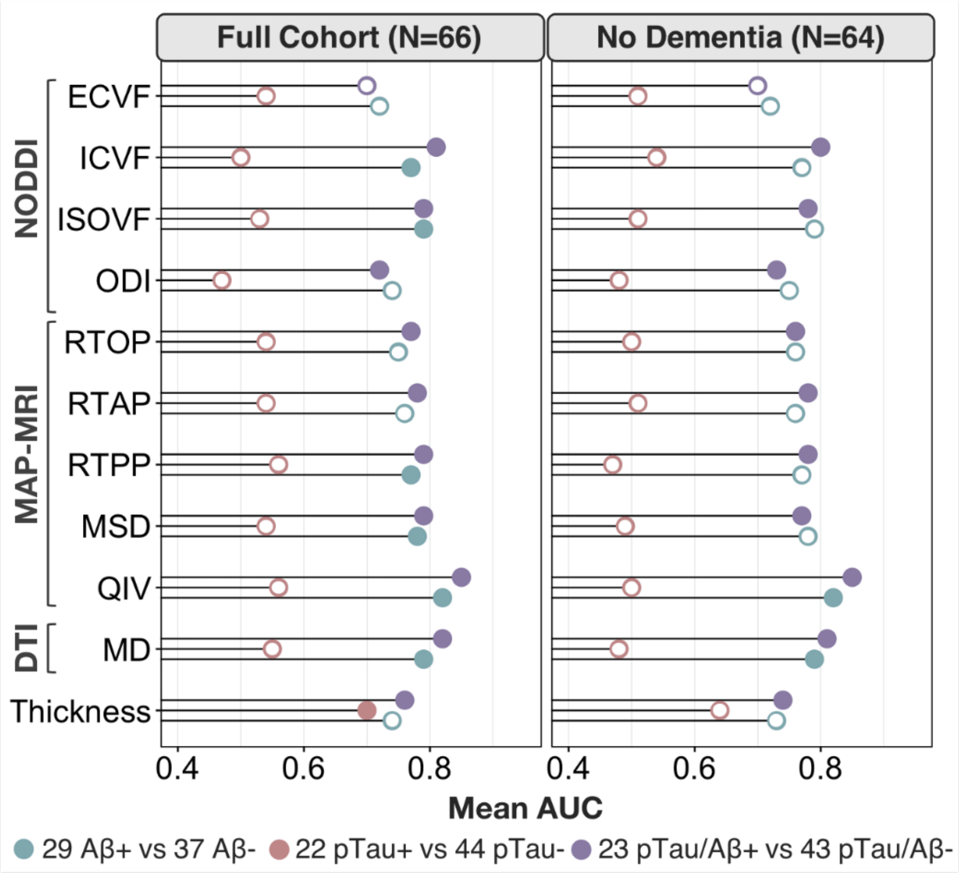

**Supplementary Figure 14.** For each residualized cortical AD-metaROI measure, the area under the curve (AUC) of the dichotomized CSF group classifications are reported. Significant associations after multiple comparisons correction are indicated by filled circles (Full cohort: dMRI $p \leq 0.020$, CTh $p \leq 0.035$; No Dementia: dMRI $p \leq 0.017$, CTh $p \leq 0.17$).

**Supplementary Table 6.** For each residualized cortical AD-metaROI measure, dichotomized CSF biomarker classification area under the curve (AUC)

| Model | Measure | Full Cohort (N=66) | | | Exclude Dementia (N=64) | | |
|---|---|---|---|---|---|---|---|
| | | **Aβ** | **pTau** | **pTau/Aβ** | **Aβ** | **pTau** | **pTau/Aβ** |
| **MT-NODDI** | **ECVF** | 0.72 | 0.54 | 0.70 | 0.72 | 0.51 | 0.70 |
| | **ICVF** | 0.77** | 0.50 | 0.81** | 0.77* | 0.54 | 0.80** |
| | **ISOVF** | 0.79** | 0.53 | 0.79** | 0.79* | 0.51 | 0.78** |
| | **ODI** | 0.74* | 0.47 | 0.72** | 0.75* | 0.48 | 0.73** |
| **MAP-MRI** | **RTOP** | 0.75* | 0.54 | 0.77** | 0.76 | 0.50 | 0.76** |
| | **RTAP** | 0.76* | 0.54 | 0.78** | 0.76* | 0.51 | 0.78** |
| | **RTPP** | 0.77** | 0.56 | 0.79** | 0.77* | 0.47 | 0.78** |
| | **MSD** | 0.78** | 0.54 | 0.79** | 0.78* | 0.49 | 0.77** |
| | **QIV** | **0.82**** | 0.56 | **0.85**** | **0.82**** | 0.50 | **0.85**** |
| **DTI** | **MD** | 0.79** | 0.55 | 0.82** | 0.79** | 0.48 | 0.81** |
| **FreeSurfer** | **CTh** | 0.74** | **0.70**** | 0.76** | 0.73 | **0.64** | 0.74** |

** Full cohort: dMRI $p \leq 0.020$, CTh $p \leq 0.035$
**No Dementia: dMRI $p \leq 0.017$, CTh $p \leq 0.17$
* $p<0.05$

##### 2.7.2.2 Five-fold logistic regression classification 80-20 split

As reported in **Supplementary Figure 15** and **Supplementary Table 7**, residualized dMRI QIV AD-metaROI measures best distinguished Aβ+ from Aβ- participants (mean AUC=0.80). In contrast, for $pTau_{181}$ classification, cortical thickness (AUC=0.72), RTAP and RTOP (AUC=0.70-0.71) performed marginally better than other measures. $pTau_{181}/A\beta_{1–42}$ status was best classified by dMRI measures (AUC=0.82-0.85), excluding ECVF, with ICVF and QIV performing marginally better (AUC=0.85). Similar classification trends were found in the subset of participants without dementia, the most notable differences being that compared to the full group 1) measures of restriction and ODI better classified Aβ status, and 2) there was an even more notable difference in $pTau_{181}$ classification accuracy between diffusion and thickness measures, with diffusion measures performing on average marginally worse than in the full group and cortical thickness performing better (AUC=0.76).

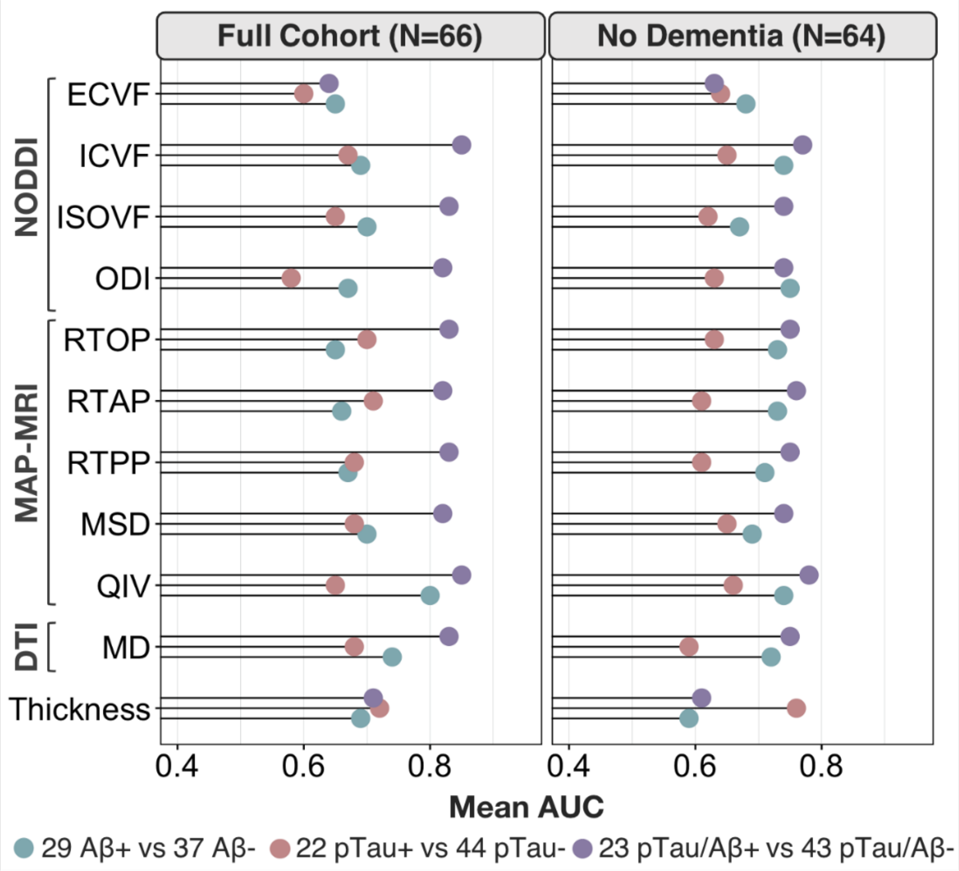

**Supplementary Figure 15.** For each residualized cortical AD-metaROI measure, the average dichotomized CSF biomarker classification area under the curve (AUC) from 5 folds is reported.

**Supplementary Table 7.** For each residualized cortical AD-metaROI measure, the average dichotomized CSF biomarker classification area under the curve (AUC) from 5 folds is reported.

| **Model** | **Measure** | **Full Cohort (N=66)** | | | **Exclude Dementia (N=64)** | | |
|---|---|---|---|---|---|---|---|
| | | **Aβ** | **pTau** | **pTau/Aβ** | **Aβ** | **pTau** | **pTau/Aβ** |
| **MT-NODDI** | **ECVF** | 0.65 | 0.60 | 0.64 | 0.68 | 0.64 | 0.63 |
| | **ICVF** | 0.69 | 0.67 | **0.85** | 0.74 | 0.65 | 0.77 |
| | **ISOVF** | 0.70 | 0.65 | 0.83 | 0.67 | 0.62 | 0.74 |
| | **ODI** | 0.67 | 0.58 | 0.82 | **0.75** | 0.63 | 0.74 |
| **MAP-MRI** | **RTOP** | 0.65 | 0.70 | 0.83 | 0.73 | 0.63 | 0.75 |
| | **RTAP** | 0.66 | 0.71 | 0.82 | 0.73 | 0.61 | 0.76 |
| | **RTPP** | 0.67 | 0.68 | 0.83 | 0.71 | 0.61 | 0.75 |
| | **MSD** | 0.70 | 0.68 | 0.82 | 0.69 | 0.65 | 0.74 |
| | **QIV** | **0.80** | 0.65 | **0.85** | 0.74 | 0.66 | **0.78** |
| **DTI** | **MD** | 0.74 | 0.68 | 0.83 | 0.72 | 0.59 | 0.75 |
| **FreeSurfer** | **CTh** | 0.69 | **0.72** | 0.71 | 0.59 | **0.76** | 0.61 |

#### 2.8 Sensitivity analysis: Mediation analyses in non-demented participants

We conducted sensitivity analyses to determine whether the 10 significant mediating effects of cortical measures on the relationship between CSF biomarkers and delayed memory would hold after excluding individuals with dementia (N=2). Overall, mediation results were similar to those in the whole sample **(Supplementary Table 8)**. Specifically, lower $A\beta_{1\text{–}42}$ ($p$=0.02; $\beta$=0.27) and greater $pTau_{181}/A\beta_{1\text{–}42}$ ($p$=5.6x$10^{-4}$; $\beta$ =-0.43) were associated with worse delayed memory performance. As in the full group, the effect of $A\beta_{1\text{–}42}$ on delayed memory were fully mediated by ICVF and QIV, while the effect of $pTau_{181}/A\beta_{1\text{–}42}$ was partially mediated by all eight dMRI AD-metaROI measures tested.

**Supplementary Table 8.** Sensitivity mediation analyses in participants without dementia.

| **Memory (DV)** | **CSF (IV)** | **MRI Model** | **Cortical Mediator** | **Direct Effect** | | **Mediation Effect** | | |
|---|---|---|---|---|---|---|---|---|
| | | | | **β** | **P** | **β** | **P** | **%** |
| **WMS Logical Memory** (N=64) | **Aβ** $p$=0.023 $\beta$=0.27 | MT-NODDI | ICVF | 0.19 | 0.098 | 0.083 | **0.034**** | 30.1 |
| | | MAP-MRI | QIV | 0.17 | 0.16 | 0.097 | **0.016**** | 36.2 |
| | **pTau/Aβ** $p$=5.6x$10^{-4}$ $\beta$=-0.43 | MT-NODDI | ICVF | -0.35 | **0.004**** | -0.09 | **0.048**** | 20.2 |
| | | | ISOVF | -0.33 | **0.008**** | -0.10 | **0.026**** | 23.8 |
| | | MAP-MRI | MSD | -0.34 | **0.006**** | -0.09 | **0.026**** | 24.6 |
| | | | QIV | -0.30 | **0.012**** | -0.10 | **0.028**** | 24.8 |
| | | | RTAP | -0.35 | **0.004**** | -0.09 | **0.028**** | 25.2 |
| | | | RTOP | -0.36 | **0.002**** | -0.08 | **0.044**** | 23.7 |
| | | | RTPP | -0.34 | **0.006**** | -0.09 | **0.038**** | 22.6 |
| | | DTI | MD | -0.33 | **0.01**** | -0.10 | **0.022**** | 24.4 |

**Indirect $p$ < FDR Threshold for 10 tests; Direct $p \leq 0.012$; Mediation $p \leq 0.048$

#### 2.9 Spatially distinct patterns of ICVF and ISOVF associations with amyloid

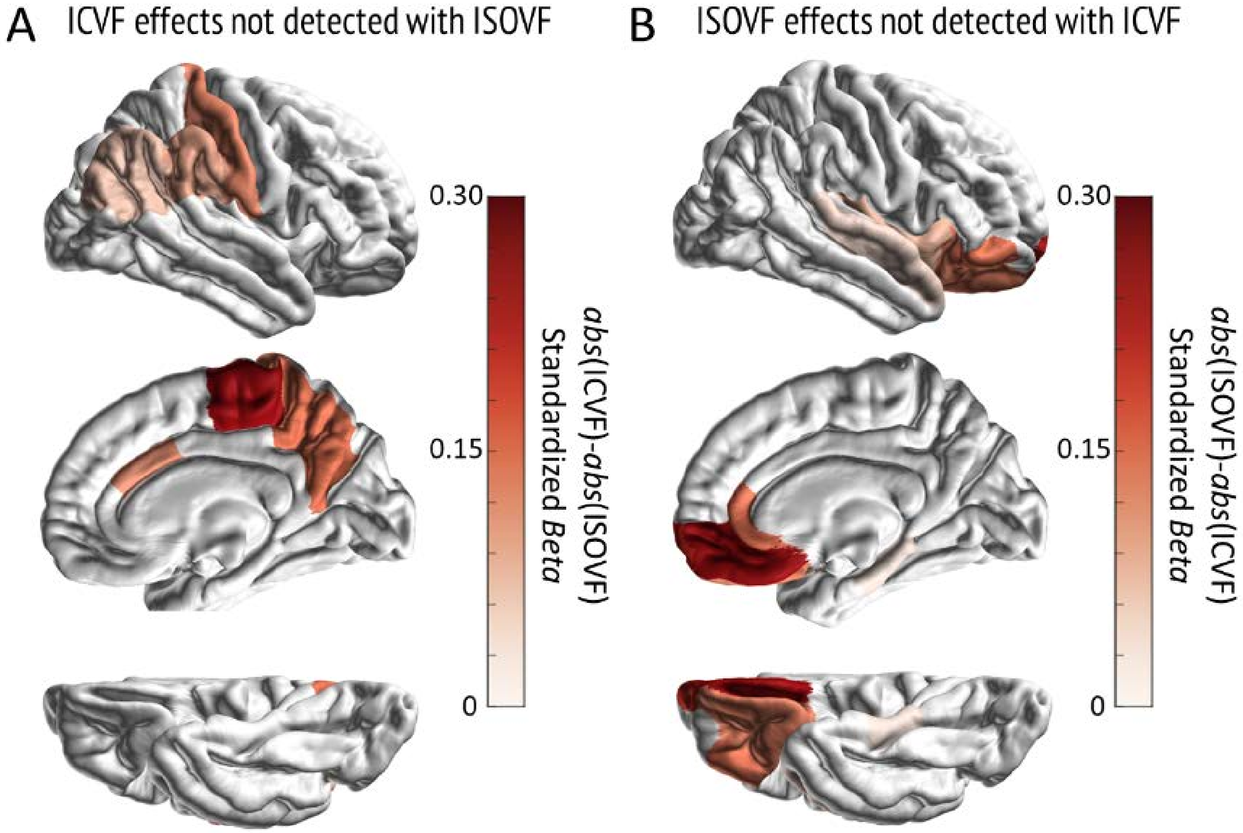

**Supplementary Figure 16.** In all regions that were significantly associated with amyloid for either ICVF or ISOVF, but not both, we took the difference in magnitude of the effect sizes to capture the diverging patterns of effects. (**A**) In regions where only ICVF associations were significant, the absolute value of the ICVF effect minus the absolute value of the ISOVF effect revealed the largest differences in early/highly myelinated sensory-motor cortical regions like the paracentral cortex. (**B**) In regions where only ISOVF associations were significant, absolute ISOVF minus absolute ICVF effects revealed the largest differences in late/lightly myelinated cortical regions like the medial orbitofrontal cortex. These dMRI measures may capture the proposed inverse relationship between the pattern of pathology progression and myelination during development (Bartzokis et al. 2007; Braak et al 1996).

#### 2.10 CSF pTau and delayed working memory

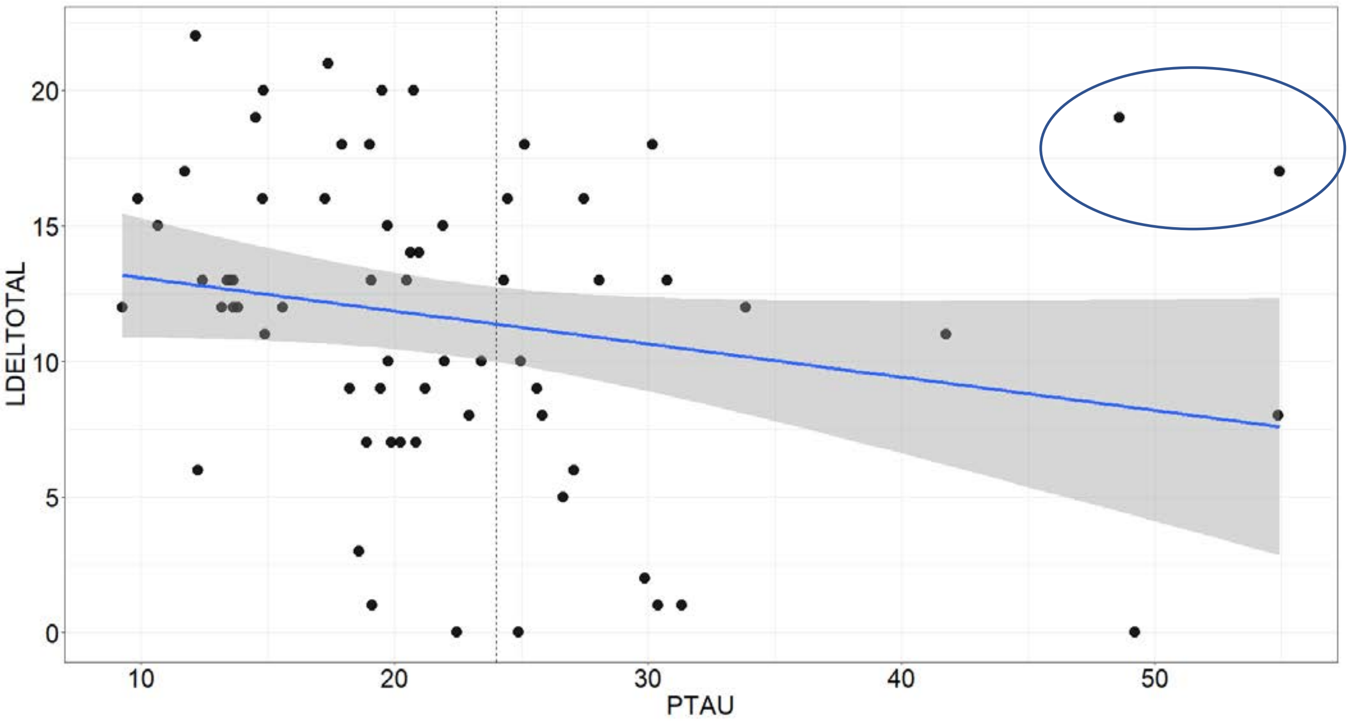

**Supplementary Figure 17.** The lack of significant associations between CSF pTau and delayed memory may be due in part to the limited number of participants in our study, particularly those with high pTau concentrations. Two CU individuals (one Aβ+ and one Aβ-) with high delayed memory scores and high pTau were sufficient to drive the association (or lack thereof).
